# A conserved role for ALG10/ALG10B and the *N*-glycosylation pathway in the sleep-epilepsy axis

**DOI:** 10.1101/2024.12.11.24318624

**Authors:** Shubhroz Gill, Torrey R. Mandigo, Ayse Deniz Elmali, Brittany S. Leger, Bo Yang, Steven Tran, Kanjana Laosuntisuk, Jacqueline M. Lane, Dalton Bannister, Chanat Aonbangkhen, Kiel G. Ormerod, Belinda Mahama, Kelsey N. Schuch, Carolyn Elya, Jamilla Akhund-Zade, Suraj R. Math, Nicholas C. LoRocco, Soobin Seo, Matthew Maher, Oguz Kanca, Nerses Bebek, Derya Karadeniz, Gulcin Benbir Senel, Carolina Courage, Anna-Elina Lehesjoki, John W. Winkelman, Hugo J. Bellen, Benjamin de Bivort, Anne C. Hart, J. Troy Littleton, Betul Baykan, Colleen J. Doherty, Girish C. Melkani, David A. Prober, Christina M. Woo, Richa Saxena, Stuart L. Schreiber, James A. Walker

**Affiliations:** Broad Institute, 415 Main Street, Cambridge, MA, USA; Center for Genomic Medicine, Massachusetts General Hospital, Boston, MA 02114, USA; Department of Neurology, Istanbul Faculty of Medicine, Istanbul University, Turkey; Department of Chemistry and Chemical Biology, Harvard University, Cambridge, MA, USA; Division of Biology and Biological Engineering, Tianqiao and Chrissy Chen Institute for Neuroscience, California Institute of Technology, Pasadena, CA, USA; Department of Molecular and Structural Biochemistry, North Carolina State University, Raleigh, NC, 27695, USA; Department of Pathology, Division of Molecular and Cellular Pathology, School of Medicine, The University of Alabama at Birmingham, AL 35294, USA; The Picower Institute for Learning and Memory, Department of Brain and Cognitive Sciences, Massachusetts Institute of Technology, Cambridge, MA, USA; Carney Institute for Brain Science, Department of Neuroscience, Brown University, Providence, RI, USA; Department of Molecular and Cellular Biology, Harvard University, Cambridge, MA, USA; Center for Brain Science and Department of Organismic and Evolutionary Biology, Harvard University, Cambridge, MA, USA; Departments of Molecular and Human Genetics and Neuroscience, Neurological Research Institute at Texas Children’s Hospital, Baylor College of Medicine, Houston Texas 77030, USA; Sleep and Disorders Unit, Department of Neurology, Cerrahpasa Faculty of Medicine, Istanbul University-Cerrahpasa, Istanbul, Turkey; Folkhälsan Research Center, Helsinki and Department of Medical and Clinical Genetics, University of Helsinki, Finland; Departments of Psychiatry and Neurology, Massachusetts General Hospital, Harvard Medical School, Boston, MA, USA; Department of Neurology, Istanbul Faculty of Medicine, Istanbul University and Department of Neurology, EMAR Medical Center, Istanbul, Turkey; Center for Genomic Medicine, Massachusetts General Hospital, Boston, MA 02114, USA; Department of Anesthesia, Critical Care and Pain Medicine, Massachusetts General Hospital and Harvard Medical School, Boston, MA 02114; Center for Genomic Medicine, Massachusetts General Hospital, Boston, MA 02114, USA; Department of Neurology, Massachusetts General Hospital and Harvard Medical School, Boston, MA 02114, USA

**Keywords:** Drosophila, human disease, glycosylation, sleep, epilepsy, neurological symptoms, cardiovascular symptoms, congenital disorders of glycosylation, glycoproteomics

## Abstract

Congenital disorders of glycosylation (CDG) comprise a class of inborn errors of metabolism resulting from pathogenic variants in genes coding for enzymes involved in the asparagine-linked glycosylation of proteins. Unexpectedly to date, no CDG has been described for *ALG10*, encoding the alpha-1,2-glucosyltransferase catalyzing the final step of lipid-linked oligosaccharide biosynthesis. Genome-wide association studies (GWAS) of human traits in the UK Biobank revealed significant SNP associations with short sleep duration, reduced napping frequency, later sleep timing and evening diurnal preference as well as cardiac traits at a genomic locus containing a pair of paralogous enzymes *ALG10* and *ALG10B*. Modeling Alg10 loss in *Drosophila,* we identify an essential role for the *N*-glycosylation pathway in maintaining appropriate neuronal firing activity, healthy sleep, preventing seizures, and cardiovascular homeostasis. We further confirm the broader relevance of neurological findings associated with Alg10 from humans and flies using zebrafish and nematodes and demonstrate conserved biochemical roles for *N*-glycosylation in *Arabidopsis*. We report a human subject homozygous for variants in both *ALG10* and *ALG10B* arising from a consanguineous marriage, with epilepsy, brain atrophy, and sleep abnormalities as predicted by the fly phenotype. Quantitative glycoproteomic analysis in our *Drosophila* model identifies potential key molecular targets for neurological symptoms of CDGs.

## INTRODUCTION

Protein glycosylation is the covalent conjugation of a glycan to the amino acids of a protein, either co- or post-translationally. This process is widespread across eukarya and archaea, and it is estimated that up to half of the human proteome is glycoconjugated. Glycosylation is critically important in diverse processes such as protein folding and trafficking ^1^, and intracellular and intercellular communication. *N*-Linked glycosylation (NLG) refers to the enzymatic *en bloc* transfer of a preassembled Glc_3_Man_9_GlcNAc_2_ oligosaccharide (lipid-linked oligosaccharide, LLO) to asparagine residues of polypeptides that satisfy the sequon Asn-X-Ser/Thr where X ≠ Pro. Several NLG enzymes are themselves *N*-glycosylated to exert negative feedback control on the pathway. The preassembly of the LLO is executed by an assembly line of enzymes in the ER, which were initially identified and characterized in yeast ^2^.

In humans, loss-of-function mutations in the orthologs of these yeast enzymes result in a set of largely-incurable, inborn errors of metabolism called congenital disorders of glycosylation (CDGs) ^3,4^. Notably, while somewhat heterogeneous overall, many CDG patients exhibit shared characteristics: reduced muscle tone, seizures, cognitive impairment, and cardiovascular abnormalities. Clinically, suspected CDGs are diagnosed using biochemical assays to quantify the glycosylation of two model proteins (transferrin and ApoC-III) in blood samples and by sequencing of cataloged candidate CDG genes ^5^. The clinical value of CDG-relevant genes has been the catalyst for the discovery of a CDG for every enzyme in the NLG pathway, except DOLPP1, ALG10/ALG10B, and ALG5. Of these, DOLPP1 and ALG5 are responsible for producing early precursors in the pathway and it is possible that they are essential for life.

Sleep is behaviorally defined as a daily period of reversible organismal quiescence characterized by reduced responsiveness to external stimuli and an elevated arousal threshold, and homeostatic regulation, all of which are essential for neurological function^6^. Human genetics has been instrumental in discovering new genes contributing to human sleep/wake regulation ^7–13^, as well as for confirming the human relevance of those discovered in model organisms. Almost all of these have relied on rare (e.g., familial) variants. More recently, genome-wide association studies of sleep traits (sleep duration, daytime napping, insomnia, and excessive daytime sleepiness) ^14–19^ and chronotype (i.e., morningness/eveningness preference) ^20,21^ have led to the discovery of hundreds of trait-associated genomic loci. These associations have illuminated the causal impact of altered sleep and chronotype on a multitude of neurological, psychological, and cardiovascular outcomes. However, aside from signals at clock genes for chronotype, where the biological mechanisms are understood in much more detail from decades of work in model organisms ^22^, the vast majority of the common variants identified in sleep GWAS are located near genes of unknown mechanistic significance and effector genes are unknown. Thus, pinpointing causal genes under association peaks could identify new elements of sleep biology and uncover new mechanisms governing healthy and aberrant sleep. We reasoned that we could identify evolutionarily conserved, novel mechanisms governing sleep by screening the orthologs of candidate human GWAS effector genes in *Drosophila melanogaster* and performing follow-up functional studies. Furthermore, the fly allows us to break synteny among human genes on a haplotype and delineate the contribution of individual genes. Additionally, phenotypic assessment of gene knockdown in multiple organisms may further clarify a functional role.

Here we delineate the human genetic locus spanning the two arms in the centromere-proximal regions of chromosome 12 containing a pair of paralogous *N*-glycosylation enzymes (ALG10 and ALG10B) that are required for neuronal homeostasis. *Drosophila* modeling confirmed human sleep traits and identified a seizure phenotype. Validating our predictions from flies, we subsequently identified an individual with progressive myoclonic epilepsy having an ultra-rare genotype of homozygous pathogenic/likely pathogenic variants in both *ALG10* and *ALG10B*. Disruption of Alg10 orthologs in zebrafish and *C. elegans*, confirmed the presence of disrupted sleep observed in humans and flies. An *Arabidopsis thaliana* salt growth phenotype can be rescued by the overexpression of human ALG10 indicating that the biochemical function of the ALG10 enzymes extends to plants. Extending observations around ALG10, we provide evidence that multiple other enzymes in the *N*-glycosylation linear ‘assembly line’ pathway are involved in sleep and epilepsy in both humans and flies. Combining an unbiased proteomic and glycoproteomic strategy to uncover the proteins in the fruit fly brain that likely mediate the neurological symptoms, we provide evidence for their role in humans, and rigorously test the assumption that perturbed *N*-glycosylation alters protein abundance.

## RESULTS

### Common variants near the paralogous genes *ALG10* and *ALG10B* encoding alpha-1,2-glucosyltransferases are associated with multiple cardiovascular and sleep traits

Motivated by the observation that multiple sleep traits exhibited association signals at two pericentromeric regions on chromosome 12, one near *ALG10* and another near its paralog *ALG10B*, we catalogued human trait associations at genome-wide (p < 5x10^-8^) significance of this region from the literature (**Figure 1A; Table S1**). The majority of curated associations were with cardiovascular (14/48, 29%) or sleep/circadian (20/48, 42%) traits. The most significant cardiovascular association was with “heart rate increase or heart rate recovery (HRR10)” (p=2.6 x 10^-66^) while the top neurological association was with “chronotype (morningness/eveningness preference)” (p=2.0 x 10^-43^). Note that these do not necessarily represent independent signals but symbolize a preponderance of associations across publications and cohorts implicating the region. The *ALG10*-*ALG10B* associations were distributed pericentromerically on both chromosomal arms and these traits fell into two major classes of related traits. Of particular interest to us, self-reported traits such as ease of waking up in the morning, daytime napping, morningness, snoring, sleep duration, and objective sleep traits measured by 7-day wrist-worn actigraphy such as daily timing of highest and lowest activity (m10 and l5) are all associated with genome-wide significance (**Figure 1B and S1A-D**).

**Figure 1.**
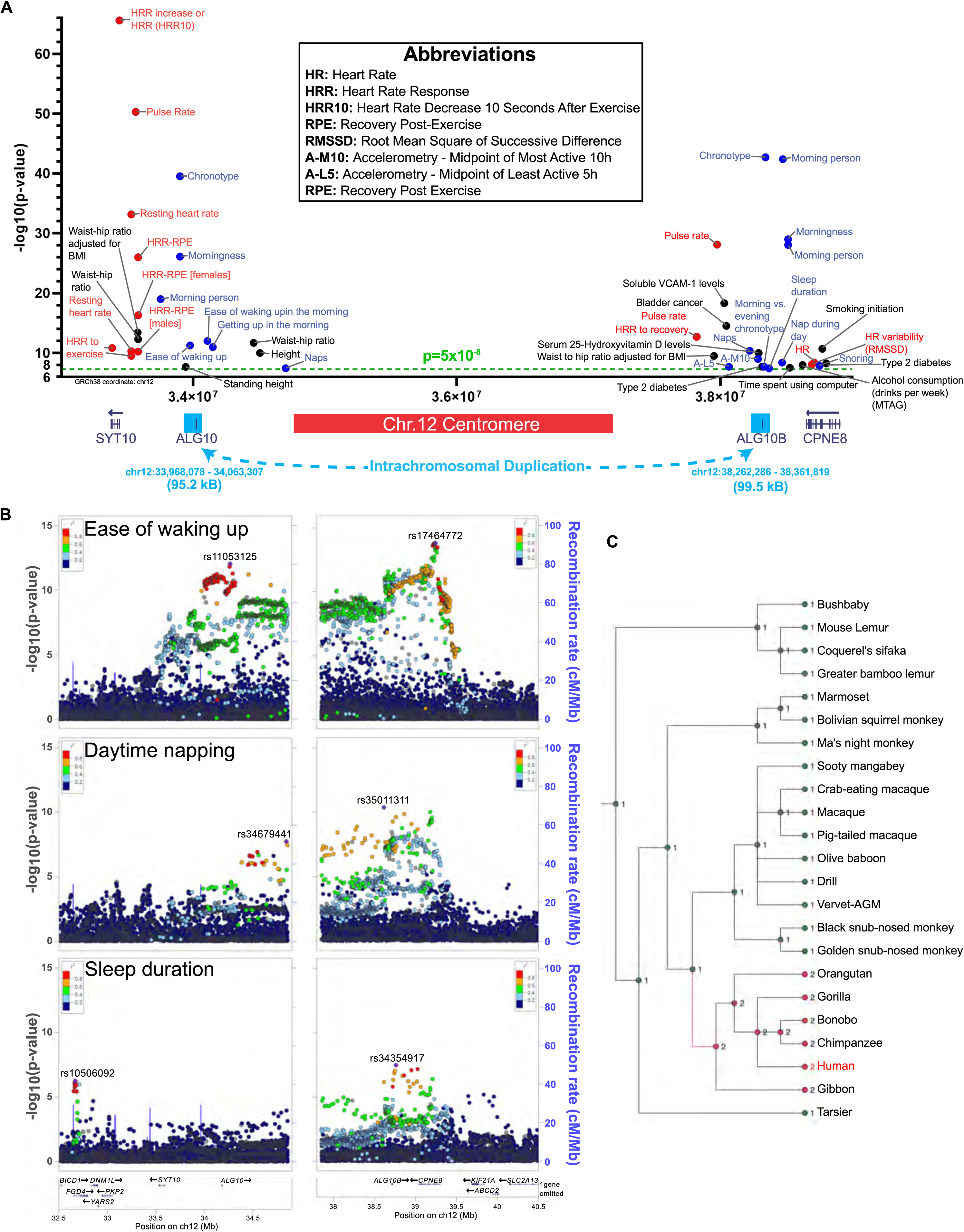
Evolutionary history and phenotypic associations in humans for the *ALG10*-*ALG10B* region. (A) GWAS associations meeting the genome-wide significance (p<5E-8, dotted green line) derived from PhenoScanner (see Methods for details) and literature in the pericentromeric region of human chromosome 12. GRCh38 coordinates are shown on the x-axis. The region encompasses the two paralogous genes *ALG10* and *ALG10B*. The region estimated to delimit the genomic region at both paralogs that was duplicated is shown below the x-axis. Each dot corresponds to a single trait in a single study. Cardiovascular traits are colored red, sleep/chronotype are colored blue, while all other traits are shown in black. (B) Regional association plots for the ALG10/ALG10B locus in GWAS of ease of waking up (n=457,776), daytime napping (n=452,633) and sleep duration (n=446,118) in the UK Biobank population. The y-axis at left shows the -log10 P value for each variant in the region and the x-axis shows the genomic position on chr 12. Each variant is represented by a filled circle, with the listed lead variant colored purple, and nearby variants colored according to correlation (r2) with the regional lead variant in the 1KG EUR population. The lower panel shows genes within the displayed region and the blue line corresponds to the recombination rate (right hand y-axis). (C) Ancestral *ALG10* gene in the primate lineage. The underlying data are derived from Genomicus version 102.01 (**Table S2**). Numbers of *ALG10*-like paralogs in each species is indicated. The speciation events in Hominoidea highlighted in red correspond to the duplication of an ancestral *ALG10*-like gene into two: *ALG10* and *ALG10B* around estimated 25 million years ago (Mya).

The associations of both *ALG10* and *ALG10B* to related traits suggested that they may share biology. Protein alignments indicated an extraordinarily high sequence conservation between ALG10 and ALG10B (95.98% identity at amino acid level, query cover = 100%, E=0.0). Furthermore, both genes are immediately adjacent to the centromere, on either side of it. Notably, centromeres are known hotspots for duplications in primates ^23–25^. Nucleotide alignments led to the identification of a ∼100kb fragment (alignment length: 102,871 bp, fraction matching: 0.9572, 95.2 kb fragment near *ALG10*, 99.5 kb fragment near *ALG10B*) that exquisitely duplicates the *ALG10*-like genes (and no other genes) creating a genetic redundancy (**Figure S1E**).

The high nucleotide and protein sequence similarity also suggested that these genes arose due to a recent duplication event. Indeed, when traced on a gene gain/loss tree of all available Ensembl genomes, a primate subclade was identified with an expansion of *ALG10* paralogs, from one ancestrally to two (**Tables S2 and S3**). We traced this event onto the primate evolutionary tree starting around 80 million years ago (Mya). Around 25 Mya, the Catarrhini clade bifurcated into Cercopithecidae (Old World Monkeys) and its sister group Hominoidea (Apes, including humans). This branchpoint was the likely stage at which the duplication event occurred, as evidenced by the presence of two *ALG10*-like genes in Apes but not in Old World Monkeys (**Figure 1C; Figure S1F**). The duplication may account for the lack of a CDG for Alg10 but needs formal verification.

### *Drosophila* modeling uncovers a role for Alg10 in sleep and seizures

Human GWAS associations at the *ALG10*-*ALG10B* locus reveal a potentially novel connection for this pair of genes to sleep and cardiovascular health. To determine whether Alg10 loss affects sleep in *Drosophila*, we used pan-neuronal (elav-Gal4) RNAi knockdown of *Alg10* and assessed locomotor activity and sleep parameters. Both elav>Alg10^RNAi^ lines (designated weak (W) and strong (S) based on knockdown efficiency determined by qPCR (**Figure S2A**)) displayed distinct features of poor quality of sleep. Flies with elav>Alg10^RNAi(W)^ showed a 14% increase in total sleep, while flies with elav>Alg10^RNAi(S)^ showed a 39% increase in total sleep (**Figure 2A; Table S4**). The increased sleep with elav>Alg10^RNAi(W)^ or elav>Alg10^RNAi(S)^ corresponded with a decrease in total activity counts (**Figure 2B)**. Investigating the characteristics of this increased sleep, we found that flies with elav>Alg10^RNAi(W)^ and elav>Alg10^RNAi(S)^ experienced 32% and 65% increases in sleep discontinuity, measured by the number of 1-minute wakes, respectively (**Figure 2C)**. Additionally, measuring the sleep of flies with elav>Alg10^RNAi(W)^ and elav>Alg10^RNAi(S)^ during daytime and nighttime sleep revealed that most of the increased sleep was during the daytime (**Figures 2D and 2E**), although there was also a significant increase in nighttime sleep with elav>Alg10^RNAi(S)^ **(Figure 2E)**. To better understand the basis of the decrease in behavioral activity, we utilized a video-based monitoring system (MARGO) **(Figure 2F)** ^26,27^. Analysis of elav>Alg10^RNAi(W)^ flies showed no difference in the speed of movement compared to controls. However, there was a significant decrease in speed of elav>Alg10^RNAi(S)^ flies compared to controls suggesting these flies may be more lethargic **(Figure 2G; Table S5)**. Additionally, in longevity experiments, the strength of Alg10 knockdown correlated with a decrease in the lifespan of elav>Alg10^RNAi(W)^ and elav>Alg10^RNAi(S)^ flies (**Figure S2B**). Since sleep and circadian rhythm are intricately connected, we subjected elav>Alg10^RNAi(S)^ flies to constant darkness to determine whether Alg10 loss affects circadian rhythm in *Drosophila*. Although there was no change in the average period or percent of rhythmic flies with elav> Alg10^RNAi(S)^ knockdown, there was a decrease in the strength of the circadian rhythm as demonstrated by a decrease in the average maximum amplitude of the Fast Fourier Transform (FFT) of the 24-hour activity profile of elav> Alg10^RNAi(S)^ flies (**Figure S3A, S3B, and S3C**). Since all previously mentioned data was gathered from male flies, the effects of Alg10 knockdown were investigated in both virgin and mated females. Compared to controls, virgin and mated elav>Alg10^RNAi(S)^ female flies experienced a 63% and 41% increase in total sleep (**Figure S3D**) which only corresponded with a significant decrease in total activity counts in virgin elav>Alg10^RNAi(S)^ females (**Figure S3E**). Unlike in male flies, elav>Alg10^RNAi(S)^ did not lead to a significant change in sleep discontinuity in virgin or mated females (**Figure S3F**). Together these data indicate Alg10 is essential for the regulation of sleep and circadian rhythm in *Drosophila* with pan-neuronal knockdown of Alg10 resulting in sleep discontinuity, increased total sleep, decreased strength of rhythmicity, and decreased lifespan.

**Figure 2.**
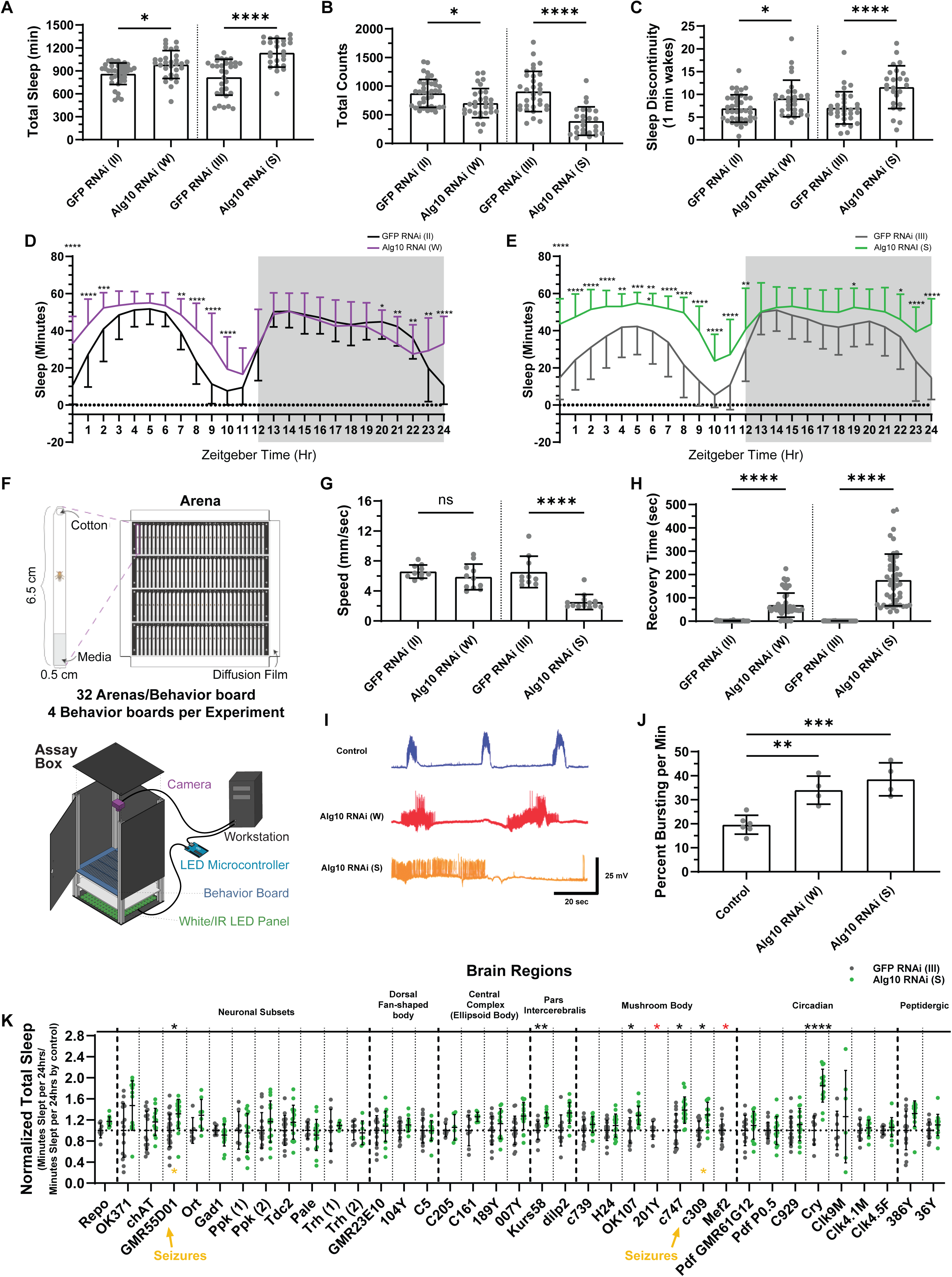
Neuronal loss of Alg10 in *Drosophila* leads to sleep defects and seizures. (A-C) Quantification of sleep parameters obtained from TriKinetics DAM system for individually housed flies. Each dot represents the mean for an individual fly over 5 days. Error bars represent the standard deviation. The indicated parameter (y-axis) for the weak (2nd chromosome) and strong (3rd chromosome) neuron-specific knockdown of *Alg10* using an elav-Gal4 line are compared to the corresponding control (GFP-RNAi) transgenic strain on the same chromosome. Adjusted p-values for the Šídák’s multiple comparisons test are indicated with asterisks. (D and E) Quantification of total sleep in hourly bins in (D) weak or (E) strong RNAi-mediated neuronal knockdown of *Alg10*. Multiple unpaired t tests with an FDR of 1% were conducted and q values are shown. (F) Schematic of MARGO (Massively Automated Real-time GUI for Object-tracking) set up for measuring locomotion of flies. (G) Locomotion speed of flies with weak (W) or strong (S) RNAi neuronal knockdown of Alg10 as determined by MARGO. Adjusted p-values from a one-way ANOVA with Šídák’s multiple comparisons test are shown. (H) Quantification of bang-sensitive seizures (recovery time from a bang-induced seizure at time t=0) in weak (W) or strong (S) knockdown of *Alg10* relative to a GFP-RNAi control on the same chromosome. Data points shown as triangles are flies that did not recover within a 480-s period and are shown at this upper limit for simplicity. Note that this implies that the indicated mean for these groups is the conservative lower limit thereof. Adjusted p-values from a one-way ANOVA with Šídák’s multiple comparisons test are indicated with asterisks. (I) Characterization of the effects of Alg10 disruption on synaptic transmission and CPG properties at the 3rd instar larval NMJ. Representative CPG recordings from 3rd instar body wall muscles of the indicated genotypes with the CNS left intact. (J) Quantification of CPG output across genotypes reveals *Alg10* knockdown larvae display a significantly greater percent of time firing compared to controls. Adjusted p-values from a one-way ANOVA with Dunnett’s multiple comparisons test are indicated with asterisks. (K) Screen of Gal4 drivers expressing within different brain regions or neuronal subtypes, to identify regions where Alg10 functions in regulating sleep. Indicated Gal4 lines were used to drive Alg10 RNAi (S). Total minutes slept was normalized to its respective control, as are the controls themselves, such that a normalized total sleep value > 1 indicates an increase in sleep compared to the control and a normalized total sleep value < 1 indicates a decrease in sleep compared to the control. Error bars represent the standard deviation. Red asterisks indicate Gal4 drivers that resulted in lethality. Yellow asterisks indicate Gal4 drivers that resulted in bang-sensitive seizures. Multiple unpaired t tests with an FDR of 1% were conducted and q values are indicated with asterisks.

Additionally, we discovered that some elav>Alg10^RNAi(W)^ and elav>Alg10^RNAi(S)^ flies experienced spontaneous seizures during routine handling. Upon further investigation, we were able to characterize these episodes as bang-sensitive seizures. We found that the strength of these seizures in elav>Alg10^RNAi(W)^ and elav>Alg10^RNAi(S)^ flies, as determined by recovery time, correlated to the strength of Alg10 knockdown **(Figure 2H; Figure S2A; Table S4)**. To characterize deficits in neuronal activity in Alg10 knockdown flies, we performed electrophysiological recordings. While the basic properties of synaptic transmission and muscle force contraction are not disrupted at the larval stage upon Alg10 knockdown (**Figure S4)**, both elav>Alg10^RNAi(W)^ and elav>Alg10^RNAi(S)^ larvae displayed enhanced excitability indicated by significant increases in the percent of time bursting per minute compared to a control line (**Figures 2I and 2J; Table S6**). These data suggest that Alg10 is essential for central pattern generator (CPG) function in *Drosophila* and knockdown in neurons results in bang-sensitive seizures.

To understand which regions and neuronal subtypes might contribute to the sleep defects and seizures with Alg10 deficiency, we screened a panel of candidate Gal4 drivers to express Alg10^RNAi(S)^. For ten of these Gal4 lines, knocking down *Alg10* resulted in a significant increase in sleep duration compared to the respective control: GMR55D0-Gal4 (cholinergic neurons), Kurs58-Gal4 (subset of the pars intercerebralis), OK-107-Gal4 (all lobes of mushroom body), c747-Gal4 (mushroom body), c309-Gal4 (all lobes of mushroom body), Cry-Gal4 (Pdf negative sLNv neurons, LN Pdf neurons, DN1 neurons). Furthermore, two of the Gal4 lines tested resulted in bang-sensitive seizures (GMR55D01-Gal4 and c309-Gal4), while two other Gal4 lines, 201Y-Gal4 and Mef2-Gal4, expressing primarily in the mushroom bodies (α-, β-, γ-lobes) resulted in lethality (**Figure 2K; Table S7**).

### Discovery of a patient with an ultra-rare human CDG-like phenotype defined by genetic variation in paralogs *ALG10* and *ALG10B*, as predicted by modeling in *Drosophila*

Our *Drosophila* experiments hinted that the absence of a human CDG associated with *ALG10* or *ALG10B* pathogenic variants might be due to functional redundancy. Based on our observations in *Drosophila*, we hypothesized that loss of function of both copies of *ALG10* and *ALG10B* might result in epilepsy. We first searched gnomAD (v2.1.1; ^28^) to understand the frequency of loss-of-function (LOF) mutations in *ALG10* or *ALG10B* alone. In support of functional redundancy, canonical transcripts for both *ALG10* and *ALG10B* were tolerant of LOF mutations (pLI = 0, upper bound fraction of o/e ratio >1). The most common predicted LOF allele in *ALG10* (c.370-1G>A, a splice acceptor site mutation) is present at Minor Allele Frequency (MAF) of 0.058% (and no homozygotes, N= 78,735 in gnomAD). The most common predicted LOF allele in *ALG10B* (p.Ser465ArgfsTer23, a frameshift) meeting the quality control criteria is present at MAF 0.089% (and no homozygotes, N=140,350 in gnomAD). In fact, no predicted LOF variants (meeting QC criteria) in *ALG10* or *ALG10B* are detected in homozygosity **(Table S8, S9)**. The chance of observing a quadruple predicted-LOF human, or a compound heterozygote for LOF alleles in both genes in this healthy cohort is vanishingly small. We thus turned our attention to disease cohorts.

Searches of neurological disease cohorts uncovered a report ^29^ of an individual from a consanguineous family exome sequenced in a cohort of patients diagnosed with progressive myoclonic epilepsy (PME) who harbored a homozygous frameshift variant in *ALG10* (p.Lys391Val*fs*\*35) generating predicted loss-of-function alleles (**Figure 3A**). Interestingly, as suggested by our *Drosophila* modeling, the patient also had a homozygous rare missense variant in *ALG10B* (p.(Leu253Trp) with an attenuated effect to complement glycosylation in a yeast assay compared to the human wild-type ALG10B^30^. We confirmed by Sanger sequencing the missense p.Leu253Trp (L253W) variant in *ALG10B* **(Figure S5).** While the affected proband is homozygous for both variants in *ALG10* and *ALG10B*, the parents and her sibling (who are all unaffected) are heterozygous. No other potentially causative variants in candidate genes were identified. In gnomAD, the L253W variant is found in generally healthy subjects at MAF ∼0.5% and there are five homozygous L253W individuals in gnomAD, which was the basis to rule out this potential causal variant for her diagnosis in the original study ^29^. The proband was born in a family with multiple rounds of consanguineous unions resulting in her ultra-rare genotype. To our knowledge, she is the only documented person with homozygous variants in both genes *ALG10* and *ALG10B*. Together with the recent duplication in primate evolutionary history, this case provides evidence of the genetic redundancy of *ALG10* and *ALG10B* and provides a possible explanation for the lack of a documented ALG10- or ALG10B-CDG case.

**Figure 3.**
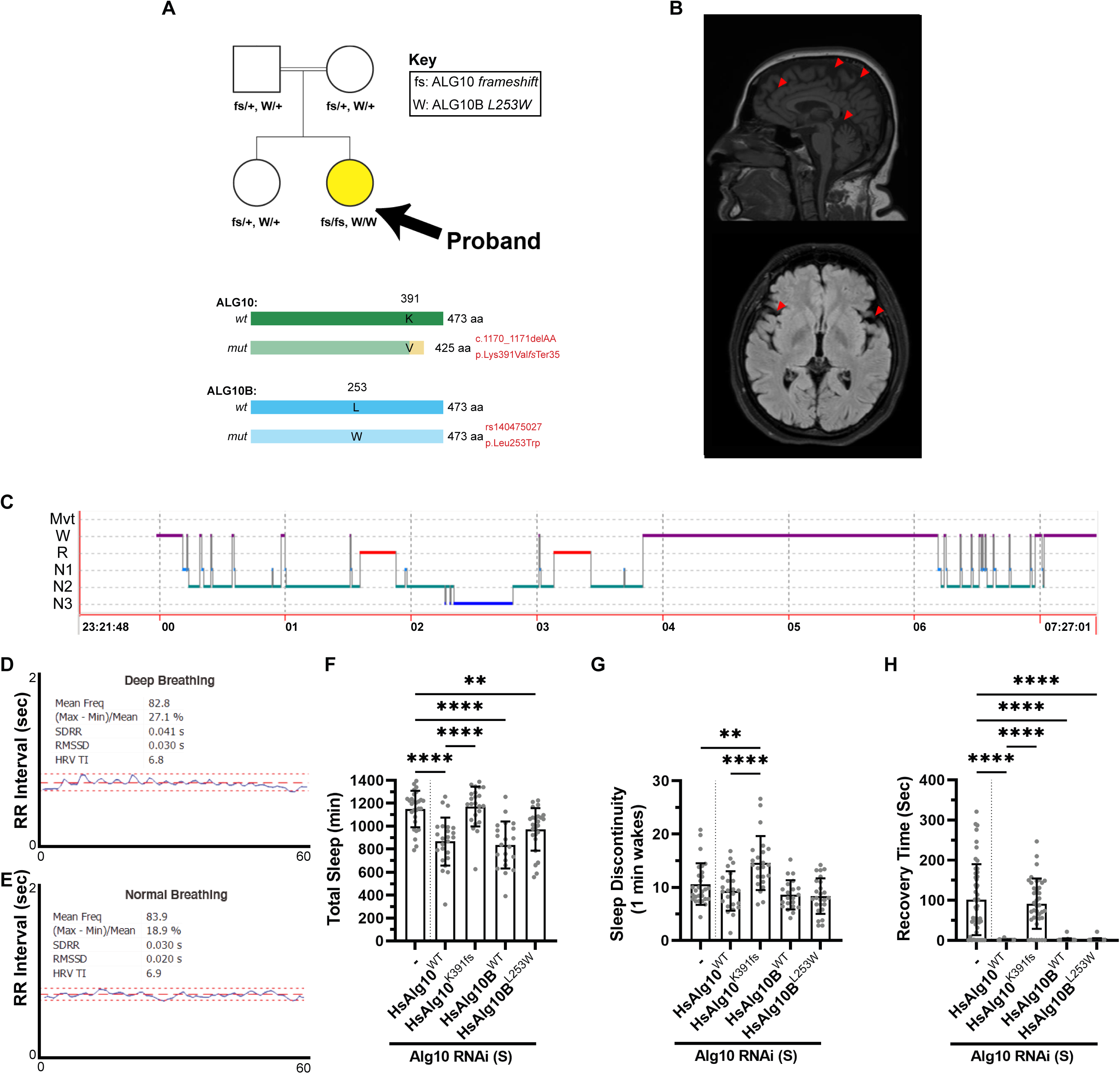
A human with an ultra-rare quadruple mutant genotype *ALG10-ALG10B* loss has epilepsy and sleep defects. (A) Partial pedigree of consanguineous union highlighting the proband (black arrow, yellow circle) with two homozygous variants: a frameshift variant in *ALG10* (ALG10 p.Lys391Val*fs*\*35) and a missense variant in *ALG10B* (ALG10B p.Leu253Trp). Inset: representation of the consequences of the two variants in the proband on ALG10 and ALG10B proteins. The proband’s parents and female sibling, who are all unaffected, are doubly heterozygous for both variants. Full pedigree covered as per medRxiv rules please contact authors for the further information. (B) MRI images highlighting the neural atrophy in the proband (red arrows highlight sites of evident regional atrophy and degeneration). (C) Hypnogram with sleep manually scored according to the AASM 2018 criteria based on overnight polysomnography from the subject during spontaneous sleep. The recording time (TRT) was 448.2 minutes, and the patient slept (TST) for 255.5 minutes. Latency to sleep onset was 12 minutes and latency to REM onset was 85 minutes. Sleep efficiency was 57%. The patient spent 155.5 minutes awake (37.8%), 14.5 minutes in N1 (3.5%), 177 minutes in N2 (43.1%), 29 minutes in N3 (7.1%) and 35 minutes in REM (8.5%). (D and E) Electrocardiogram (EKG) of the proband indicated an R-R interval, normal for age. (F and G) Quantification of total sleep and sleep discontinuity in *Drosophila* elav>Alg10 RNAi (S) rescued with transgenic expression of human wild-type ALG10, ALG10 with patient frameshift, or wild-type ALG10B. Adjusted p-values from a one-way ANOVA with Šídák’s multiple comparisons test are shown. (H) Rescue of Alg10 RNAi bang-sensitive seizures with human ALG10, ALG10 with patient frameshift, and ALG10B. P-values shown are for comparisons of the rescue construct to the respective no-human-transgene condition. Adjusted p-values from a one-way ANOVA with Šídák’s multiple comparisons test are shown.

At the time of the recall, the proband was a female in their 30’s. Her initial symptom was myoclonic jerks in her extremities, induced mainly by awakening, movement, and stress, which had started in her teens. An ataxic gait became prominent concurrently with a mild progression rate. Several years later, she had two generalized tonic-clonic seizures during her sleep. Valproic acid treatment (1000 mg/day) led to resolution of the generalized seizures and minimized the myoclonic jerks, however it had to be discontinued due to side effects, including prominent weight gain. She is currently using a combination of levetiracetam (3000 mg/day), zonisamide (100 mg/day) and clonazepam (0.5 mg/day) and has been seizure free other than jerks induced by stressful situations. She has mild cognitive dysfunction shown by neuropsychological testing, scoliosis, and reactive depressive episodes. Her cranial magnetic resonance imaging revealed cortical atrophy and prominent cerebellar atrophy (**Figure 3B**). Her electroencephalographs showed fast rhythmic activity of low amplitude along with rare theta waves. She also showed type 1 photosensitivity during 5 Hz photic stimulation ^31^ in her 30’s. Her detailed cardiologic examination and electrocardiograms (ECGs) were entirely normal, thus excluding QT prolongation. Her RR interval variation study was interpreted as normal (**Figure 3E**).

Although she had not previously been diagnosed with any sleep issues, we postulated based on our findings in the human GWAS and fly modeling that she may have an undiagnosed sleep disorder. An interview with the subject revealed her persistent difficulties with sleep. The patient was referred to the Sleep and Disorders Unit, Cerrahpasa Faculty of Medicine, Istanbul University for both the evaluation of sleep macro- and micro-organization and to investigate the presence of any sleep disorder using polysomnography (PSG) and actigraphy (**Figure 3C and 3D).** At the time of overnight PSG, the patient took the medications levetiracetam (1500 mg morning and evening, zonisamide (200 mg in the morning) and clonazepam (2 mg in the evening). The patient had significant difficulty maintaining sleep with a prolonged awakening from 4-6am and limited sleep thereafter. An excess of both alpha and sleep spindles was observed in N2 and N3 sleep, likely related to the clonazepam. Mild, REM-predominant, sleep apnea (apnea-hypopnea index (AHI) = 9/hour with REM AHI = 36/hour) was observed with mild oxygen desaturation (**Figure 3C**). Actigraphy (>1 week) not concurrent with PSG showed significantly longer sleep onset latency and lower sleep efficiency in the patient as compared to her sister, mother and father (**Table S10**).

To further investigate the functional impact of the *ALG10* frameshift and *ALG10B^L^*^253^*^W^*variants on sleep and epilepsy, we expressed human ALG10 or ALG10B from transgenes in neurons in elav>Alg10^RNAi(S)^ flies. While expression of wild-type human ALG10, ALG10B, or mutant *ALG10B^L^*^253^*^W^* rescued both the sleep deficits and bang-sensitive seizures, the *ALG10* frameshift failed to rescue these phenotypes and experienced 37% more 1-minute wakes than elav>Alg10^RNAi(S)^ controls (**Figures 3E, 3F and 3G)**. These findings not only support the conserved role of ALG10, but also demonstrate the nature of the alleles, with the *ALG10* frameshift being a strong LOF, while the *ALG10B^L^*^253^*^W^*is more likely a hypomorphic mutation since it did not rescue to as full an extent as wild-type human ALG10B. *In silico* prediction carried out by PolyPhen2 ^32^ and SIFT ^33^ assigned the variant as “possibly damaging” and “deleterious” respectively.

### Modeling Alg10 loss reveals a functionally conserved role across animal and plant kingdoms

Our results in humans and *Drosophila* suggested that alpha-1,2-glucosyltransferases may be key enzymes governing neurological traits. To understand the extent of this functional conservation, we validated our findings in three other model organisms: *Danio rerio*, *C.elegans*, and *Arabidopsis thaliana* (**Figure 4A**). Our motivation for using the first two was their well-established role in studying sleep, while the latter was to understand whether conservation of enzyme activity is restricted to the animal kingdom or whether it might extend to plants.

**Figure 4.**
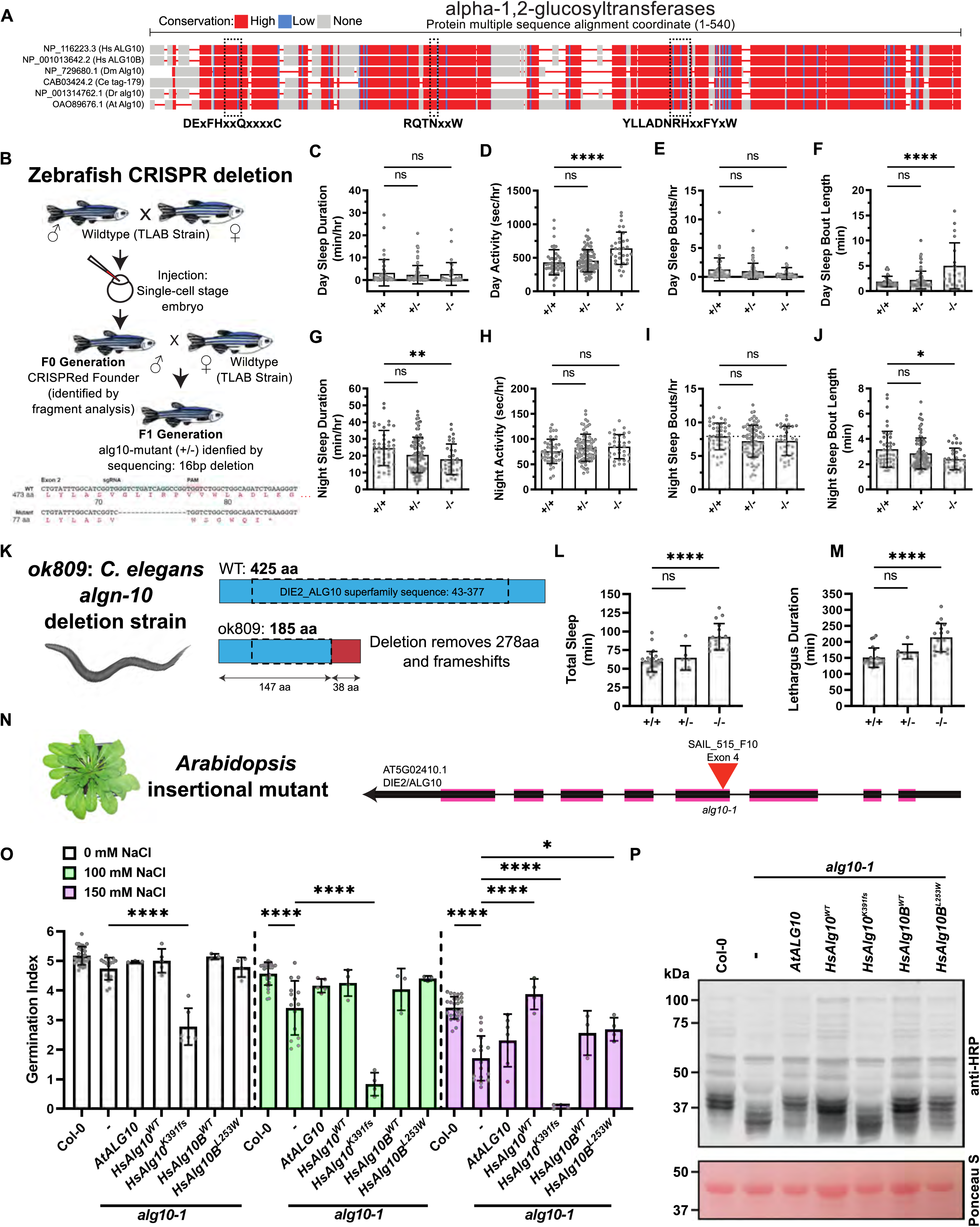
An evolutionarily conserved role for Alg10 in sleep in animals and functional conservation in plants. (A) Alignment of ALG10-like proteins from human (Hs), fruit fly (Dm), nematode (Ce), zebrafish (Dr), and Arabidopsis (At). Three sets of conserved amino acid stretches that potentially correspond to catalytic residues of the enzymes are highlighted. (B) Schematic for the generation of a zebrafish *alg10* mutant using CRISPR/Cas9, resulting in a predicted truncated 77-amino acid mutant protein. (C-F) Quantification of daytime and (G-J) nighttime sleep parameters in 6 dpf zebrafish. Each dot represents a single larva and a single day of observation. Mean +/- SD is graphed. Adjusted p-values from a one-way ANOVA with Dunnett’s multiple comparisons test are shown. (K) *C. elegans ok809* strain with deletion in *algn-10* (also known as *tag-179*) results in predicted truncated 278 amino acid protein. (L and M) Quantification of sleep in *C. elegans algn-10* (*ok809*) deletion strain (*ok809*). Each dot represents a single worm. Adjusted p-values from a one-way ANOVA with Dunnett’s multiple comparisons test are shown. (N) Thale cress (*Arabidopsis thaliana*) *alg10-1* insertional mutant showing the *DIE2*/*ALG10* locus with red triangle indicates position of T-DNA insertion. (O) *alg10-1* mutant plants have significantly delayed germination under sodium chloride stress. Rescue of the *alg10-1* mutant germination defect by *AtALG10*, *HsAlg10^WT^*, *HsAlg10^K^*^371f^*^s^*, *HsAlg10B^WT^*, and *HsALG10B^L^*^253^*^W^* under the control of a ubiquitous 35s promoter. Adjusted p-values from a one-way ANOVA with Šídák’s multiple comparisons test are shown. (P) Immunoblot analysis of total protein crude extracts from Col-0, *alg10-1*, and overexpression of *35s::AtALG10*, *35s::HsAlg10^WT^*, *35s::HsAlg10^K^*^371f^*^s^*, *35s::HsAlg10B^WT^*, and *35s:: HsALG10B^L^*^253^*^W^* in *alg10-1* mutant using anti-HRP. Proteins were separated on 7.5% SDS-PAGE, and the blot was stained with Ponceau S solution prior to immunodetection.

CRISPR/Cas9 genome editing was used to mutate the single zebrafish *alg10* ortholog (**Figure 4B**). Animals carrying a germline *alg10* mutation were identified, which consisted of a 16 bp deletion that results in a translational frameshift and predicted to generate a truncated non-functional protein (77 compared to 473 amino acids in wild-type). Sleep and locomotor activity were quantified using an automated video tracking system ^34^ to compare the behavior of homozygous mutant, heterozygous mutant, and wild-type siblings (**Figures 4C-4J; Table S11**). Homozygous *alg10* mutant fish slept 27% less at night compared to their wild-type siblings, due to a 24% decrease in nighttime sleep bout length (**Figures 4G and 4J**). Although, no significant differences in sleep duration were observed during the day, homozygous *alg10* mutant fish exhibited a 168% increase in daytime sleep bout length (**Figures 4C and 4F**). Homozygous *alg10* mutant fish also displayed a 48% increase in daytime waking activity compared to their wild-type siblings (**Figure 4D**).

To understand the role of *algn-10* (also known as *tag-179* or *T24D1.4*) in nematodes, we used the deletion allele *ok809* that was previously characterized as loss-of-function ^35^ (**Figure 4K**). Total sleep in *algn-10* homozygous mutant nematodes was 56% more than observed in wild-type controls, whereas the change in heterozygotes was not statistically significant (**Figure 4L; Table S12**). Lethargus duration, which is the time during which sleep bouts occur was also changed. Lethargus duration in homozygotes was 42% longer than wild-type controls, while heterozygotes showed an insignificant increase of 13% (**Figure 4M**). The increased total sleep and increased lethargus duration observed here may be a consequence of poor sleep efficiency which leads to compensatory sleep, as observed in other genotypes ^36–38^.

*Arabidopsis thaliana* has a single *ALG10* ortholog (*AtALG10*). To understand whether the role of ALG10 in glycosylation is conserved between humans and plants, we examined both plant phenotypes previously associated with *N-*glycosylation enzymes and biochemical effects. We first attempted to replicate a published leaf size phenotype ^39^ in *AtLG10* mutants but were hampered by extensive variability in this mutant based on subtle variations in light, temperature, and other environmental conditions. Seeking a more robust phenotype for *alg10-1* mutant plants (**Figure 4N; Table S13**), we focused on germination under salt stress, a phenotype observed in mutants of other plant *N*-glycosylation enzymes ^39–42^. Germination index (combining germination percentage and mean germination time metrics) with and without NaCl stress in wild-type (*Col0*) and *alg10-1* mutant plants was quantified (**Figure 4O**). We found that the germination index was significantly reduced in *alg10-1* mutant plants compared to *Col0* controls at 100 and 150 mM NaCl (**Figures 4O, S6**). Furthermore, these *alg10-1* phenotypes could be rescued by transgenic expression of either human ALG10, ALG10B or ALG10B^L253W^, while ALG10^K391fs^ failed to restore, further pointing to its loss-of-function nature (**Figure 4O**).

Finally, we investigated the biochemical consequences of loss of *AtALG10* using lysates from plants and immunoblot analysis with anti-HRP to detect mobility of *N*-glycosylated proteins (**Figures 4P, S9**). Transgenic expression of wild-type human ALG10, ALG10B, or mutant *ALG10B^L^*^253^*^W^* were able to rescue the altered mobility of HRP-detected proteins in *alg10-1* mutants. In contrast, the human ALG10 frameshift was unable to restore the *alg10-1 N*-glycosylated protein mobility phenotype in plants (**Figure 4P**), similar to the inability of this mutant to rescue *Drosophila* phenotypes.

### Modeling of other *N*-glycosylation pathway enzymes in fruit flies supports a pathway-level role in sleep, epilepsy, and cardiovascular function

Biochemical genetics in yeast show that the *N*-linked glycosylation pathway is primarily a linear enzymatic pathway with the product of each enzymatic step acting as the substrate for the subsequent enzyme ^43–46^. This model predicts that loss of any step of this process via mutation or knockdown should produce a similar end-result, namely reduction in or absence of the transfer of a pre-assembled LLO to substrate polypeptides (**Figure 5A**). We hypothesized that disruption of other enzymes in the *N*-glycosylation pathway might phenocopy Alg10 loss. To test this, we targeted additional enzymes of the *N*-glycosylation pathway using RNAi in *Drosophila* neurons. As with Alg10, knock down of Alg1, Alg14, CG3149 (RFT1) and Alg5 (wol) resulted in increased total sleep (**Figure 5B and Table S16**). Knock down of Alg14, RFT1 and Gny also lead to an increase in sleep discontinuity (**Figure 5C)**. Furthermore, knock down of Alg7, Alg14, Alg1 Alg2 and Xit resulted in susceptibility to bang-sensitive seizures (**Figure 5E and Table S16**). Together these data demonstrate a neuronal knock down phenotype continuum ranging from no effect (Dolk, Alg3, Alg9 females and Alg12), to short-lived flies or lethality (Alg7, Alg13, Alg2, Alg9 males and Xit) with abnormal sleep and seizures falling between the two extremes (**Figure 5F)**.

**Figure 5.**
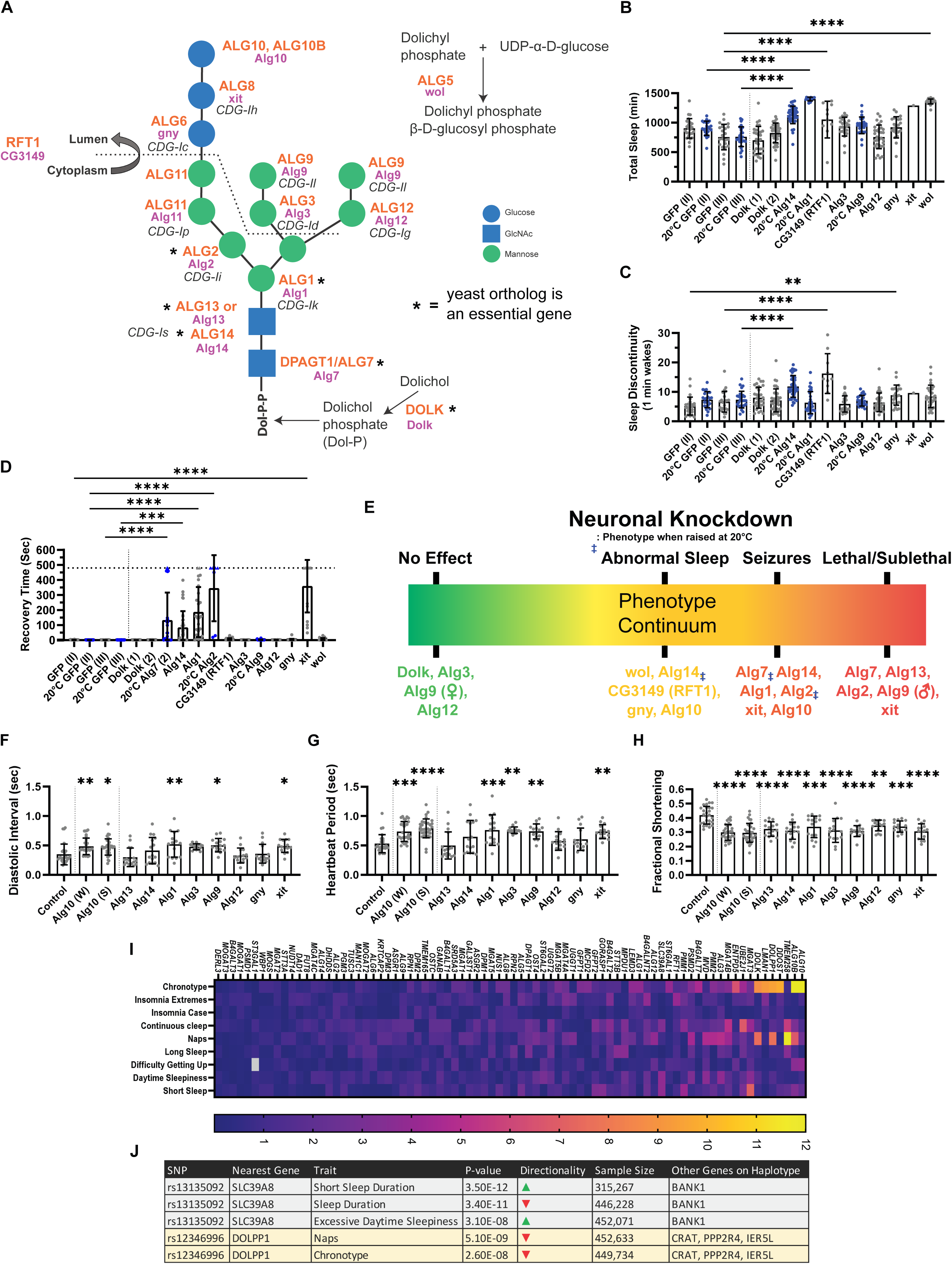
The *N*-glycosylation pathway is involved in sleep, epilepsy, and cardiovascular function in *Drosophila* and humans. (A) Overview of enzymes catalyzing the individual steps of the lipid-linked oligosaccharide (LLO) glycan. Human genes are capitalized and in orange. Orthologous fly genes are in pink under each human gene. Alg13 and Alg14 function as a complex. Asterisks mark genes whose yeast orthologs are known to be essential genes. (B and C) Sleep-related parameters in control (GFP-RNAi, RNAi transgene on the 2nd or 3rd chromosome) or gene-specific pan-neuronal (elav-Gal4) knockdown of *N*-glycosylation pathway enzymes. Adjusted p-values from a one-way ANOVA with Šídák’s multiple comparisons test are shown. (D) Seizure phenotypes (recovery time after a bang-induced seizure) in neuronal knockdown of indicated genes. Errors bars represent standard deviations. Adjusted p-values from a one-way ANOVA with Šídák’s multiple comparisons test are shown. (E) Continuum of neuronal phenotypes observed in the knockdown of *N*-glycosylation pathway enzymes. (F-H) Effect of cardiac-specific knockdown (using a Hand-Gal4 driver) of the indicated *N*-glycosylation genes in flies on (F) diastolic interval, (G) heartbeat period, and (H) cardiac contractility (fractional shortening) as assessed by videographic recordings in *ex vivo* preparations. A dotted line corresponding to the control is included to aid visualization. Adjusted p-values from a one-way ANOVA with Dunnett’s multiple comparisons test are shown. (I) Gene-based association tests of self-reported sleep trait GWAS for 34 human *N*-glycosylation enzymes. Heat-map represents p-values of gene-based association results for each gene (columns) across all traits (columns) using *PASCAL* (summing statistics across independent signals in each gene). The legend indicates lower p-values (purple to yellow). (J) Lead variants near genes encoding two *N*-glycosylation enzymes, *SLC39A8* and *DOLPP1*, are associated at genome-wide significance with sleep traits in the UK Biobank.

Based on cardiovascular trait associations with the *ALG10*-*ALG10B* locus in humans (**Figure 1A**), we investigated the role of Alg10 in *Drosophila* heart by cardiac tissue restricted knockdown using the Hand-Gal4 driver (**Figures 5F-5H**). This cardiac restricted knockdown of Alg10 resulted in a decreased lifespan with both Alg10^RNAi(W)^ and Alg10^RNAi(S)^ (**Figure S2C**). Compared to a Hand-Gal4 control, both Hand>Alg10^RNAi(W)^ and Hand>Alg10^RNAi(S)^ flies had an increased diastolic interval and heart period (**Figure 5F and 5G**). Knockdown of Alg10 also resulted in decreased fractional shortening, a dimensionless quantity that is a measure of cardiac contractility and thus health, by almost 22% in Alg10^RNAi(W)^ and 23% in Alg10^RNAi(S)^ flies as compared to the control (**Figure 5H**). To assess the concordance of *N*-glycosylation pathway phenotypes in a different tissue, we also tested the effects of heart-specific RNAi of these enzymes on cardiovascular structure and function (**Figures 5F-5H**). Once again, consistent with the observed phenotype of *Alg10*, the knockdown of eight other genes (*Alg1*, *Alg3*, *gyn*, *xit*, *Alg9*, *Alg12*, *Alg13*, and *Alg14*) led to the increased diastolic interval compared to controls (**Figure 5F**). Alterations of additional cardiac parameters linked with cardiac-specific knock-down of Alg10 RNAi and other *N*-glycosylation pathway enzymes are shown in **Table S17.** Thus, the phenotypes for the assessed *N*-glycosylation pathway were remarkably consistent in their directionality and to some extent their effect size, supporting a linear ‘assembly line’ model of *N*-glycosylation enzymes in this tissue.

We next assessed whether human genetic evidence supports a role for enzymes involved in N-glycan assembly other than *ALG10* and *ALG10B* in sleep or chronobiological traits. Common haplotypes harboring variants at or near *N*-glycosylation enzymes are associated with sleep or chronobiological traits (**Figures 5I, 5J, S9; Tables S17 and S18**). Gene-based analyses identified associations at up to 8 additional glycan synthesis enzymes with chronotype and naps of 80 genes assessed.

Together, our results in humans and flies provide multiple lines of direct and corroborating evidence that the *N*-glycosylation pathway is critical for normal brain and heart function, and that in broad consistency with the linear character of the pathway, similar phenotypes are observed upon disrupting distinct enzymes in the pathway.

### Glycoproteomics in flies uncovers potentially disease-relevant *N*-glycosylation targets of Alg10

To uncover targets of the *N*-glycosylation pathway, we performed proteome and glycoproteome profiling in adult fly brains with elav>Alg10^RNAi(S)^ and elav>Alg10^RNAi(VS)^ (an additional RNAi, derived from a VDRC GD plasmid, with very strong phenotypes) and control flies (**Figures 6A**). Our approach allowed us to detect distinct *N*-glycosylation sites (glycosites) on peptides and map these to the proteins that they originated from. The primary advantage of the glycosite-specific analysis is the higher resolution gained when detecting changes in glycan occupancy at a site level, as glycoproteins can carry multiple glycosites with distinct occupancy changes. Using this approach, we detected and quantified 152 distinct N-linked glycoproteins corresponding to 253 individual glycosites at a 5% false discovery rate (FDR) (**Figures 6B, 6C**).

**Figure 6.**
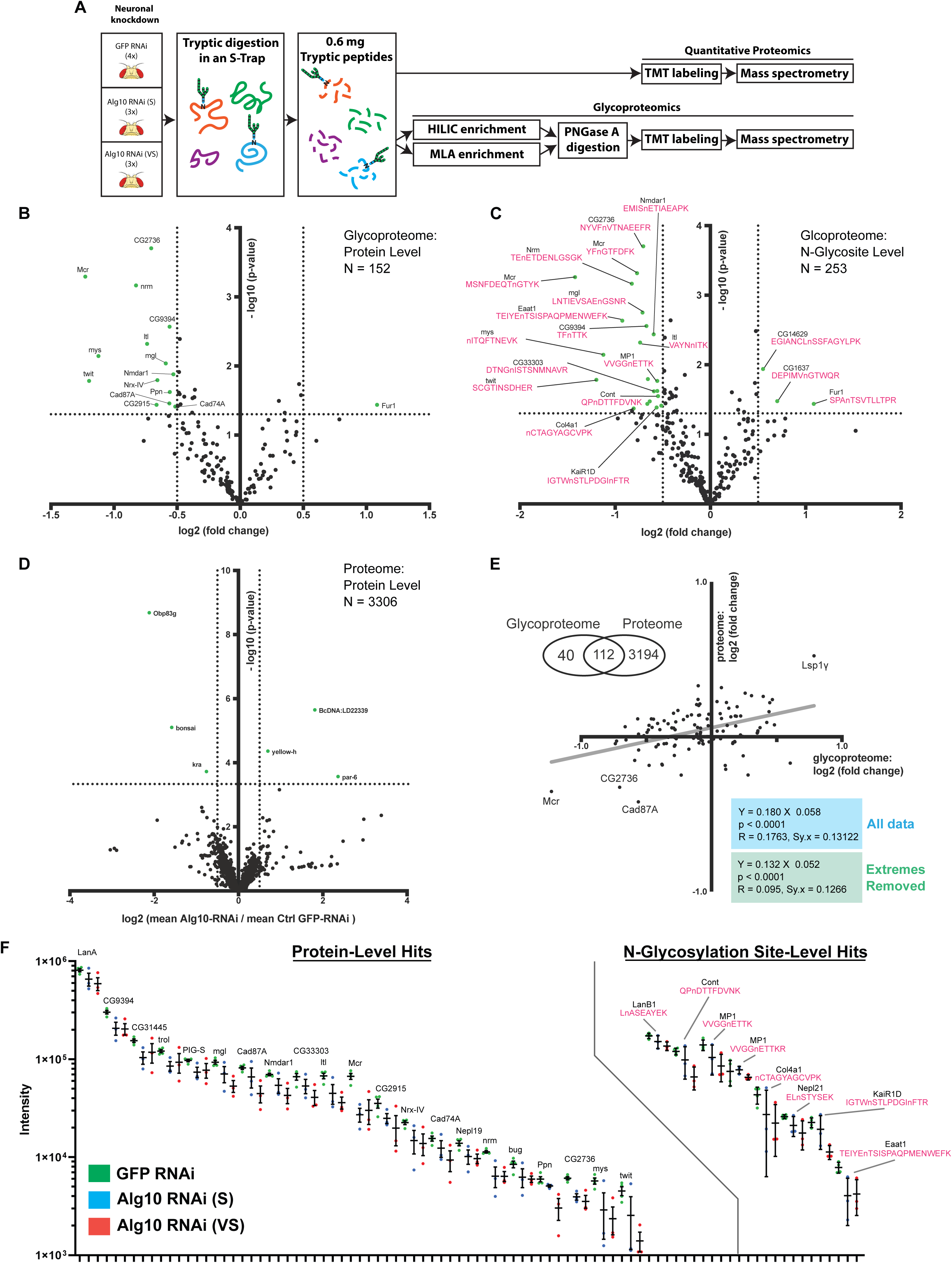
Quantitative proteomics and glycoproteomics identifies targets of Alg10 in the fly brain. A) Overview of proteomics and glycoproteomics using *Drosophila* brains. (B) Changes in the glycoproteome at the protein level (i.e., all quantified glycosylation sites in a given protein are averaged). Each dot represents one protein. Proteins that are altered (up or down) in the ALG10 RNAi brain relative to the control above the multiple-testing threshold are highlighted in green. (C) Peptide-level changes in individual *N*-glycosylation sites (“glycosites”) are shown (i.e., each glycosylation site within a given protein is treated as separate). Each dot represents one peptide. Glycopeptides that are altered (up or down) in the Alg10 RNAi brain relative to the control above the multiple-testing threshold are highlighted in green and the glycopeptide sequence itself is shown in pink. (D) Global proteomic changes in the Alg10 RNAi brain versus the control. Green dots represent changes that are considered statistically significant (p-value ≤ 0.05). (E) Comparison of glycoproteomic and proteomic changes in 112 proteins that were detected in both rounds of proteomics and glycoproteomics. (F) Quantification of individual proteins (left) or peptides (right) from control and Alg10-knockdown brains by mass spectrometry analysis in (B) and (C). A protein is only shown in the site-level set if it did not score as a protein-level hit. Note that the scale is logarithmic. The relevant peptide sequences (right half) are shown in pink.

Using a cutoff of 1.4-fold increase or decrease (log2 cutoff = ± 0.5) in overall *N*-glycosylation levels on the glycoprotein, we identified 15 out of a total of 152 proteins that were consistently altered in both RNAi lines across replicates (adj p-value ≤ 0.05). Of these, 14 (CG2736, Mcr, nrm, mgl, CG9394, Nmdar1, ltl, mys, Nrx-IV, twit, Ppn, Cad87A, CG2915, and Cad74A) displayed decreased glycosylation (‘down glycosylated’) while a single protein (Fur1) showed increased glycosylation (‘up glycosylated’) consistent with the expected directionality of effect associated with a loss of Alg10 (**Figure 6B**). When analyzed at the level of individual peptides, 31 out of a total of 253 were reproducibly altered at an adj p-value ≤ 0.05. Once again, a majority (21) of the altered glycosylation sites were down glycosylated, and only 3 were up glycosylated (**Figure 6C**). We additionally quantified a total of 3306 proteins from whole proteome analysis of adult brains (**Figure 6D**). Unlike the glycoproteome, few proteins in the proteome exhibit marked changes in either Alg10^RNAi^ condition with only 6 out of 3306 proteins had altered levels at a log2 cutoff = ± 0.5 and adj p-value ≤ 0.05. We used the 112 proteins that were detected and quantified in both the glycoproteomic and proteomic experiments to determine whether there was any association between the levels of a protein and its glycosylated form. A conservative estimate of the correlation (in the log-transformed space) after removing the two extreme points (Mcr and Lsp1γ) indicated a positive correlation (Y = 0.1322 X + 0.05216, R2 = 0.0945, p =0.001) between the log2(fold change of protein, FCP) and log2(fold change of glycoprotein, FCG) levels (**Figure 6E**). Note that in the light of the logarithmic transformation, this implies the approximate, monotonically increasing, and nonlinear relationship defined by FCP = (1.0368) x (FCG) 0.1322 between the fold changes of the protein and the glycoprotein. Visualization of the triplicates of the two RNAi lines also supports the strength of effect: in general, the glycopeptide levels in the Alg10^RNAi(S)^ condition were intermediate between those in the control and Alg10^RNAi(VS)^ brains (**Figure 6F**). Overall, these data emphasized the significant *N*-glycosylation changes affected by *ALG10* in flies.

### Genetic studies identify conserved targets of Alg10 relevant to sleep and seizures

Our proteomic-glycoproteomic profiling approach revealed that reduced *N*-glycosylation in fly neurons upon Alg10 knockdown also resulted in lowering of the target protein levels (**Figure 6E**). We speculated that reduced protein levels might be causal for the sleep and seizure Alg10^RNAi^ phenotypes. To test this, we systematically knocked down the expression of target proteins in adult neurons and determined whether this induced sleep and/or seizure phenotypes (**Table S25**).

Pan-neuronal RNAi knockdown of four tested genes - *Cad87*, *LanA*, *Nepl19*, and *Nmdar1*, each resulted in significant reduction of sleep compared to controls (**Figure 7A**). Furthermore, RNAi targeting of two genes in neurons - *CG2736* and *Nmdar1* lead to a significant increase in sleep discontinuity (**Figure 7B**). Knockdown of *CG2915*, a member of the M14 family of zinc carboxypeptidases (renamed here as *dCPA1*) led to sub-lethal flies when raised at 25°C (**Figures 7C and 7D**) which experienced long-lasting bang sensitive seizures (**Figure 7E**). Since these flies did not survive the length of the Trikinetics monitoring, knockdown of *dCPA1* was done in flies raised at 20°C. In these experiments, pan-neuronal knockdown of *dCPA1* resulted in a 32% increase in total sleep and a 37% increase in sleep discontinuity (**Figures 7F and 7G**). Together these data demonstrate that *Alg10* and *N*-glycosylation regulate multiple target proteins that are necessary for the regulation of healthy sleep. Furthermore, these data suggest that the dysregulation of dCPA1 caused by loss of Alg10, and therefore disrupted *N*-glycosylation, may be a key driver of the sleep and seizure phenotypes seen in *Alg10* knockdown flies. In addition to dCPA1, we noted that 12 other proteins identified in our fly brain glycoproteomic analyses had orthologs in humans that are linked to monogenic epilepsy (*CNTN2, GRIN1, GRIN2A, GRIN2B, GRIN2D, LAMA3, PIGS, SCARB2, SLCA12 and THSD7A*) or GWAS loci for epilepsy (*GRIK2, GRIN3A*), and 40 orthologs that lie within GWAS loci for sleep disorders (**Table S26**).

**Figure 7.**
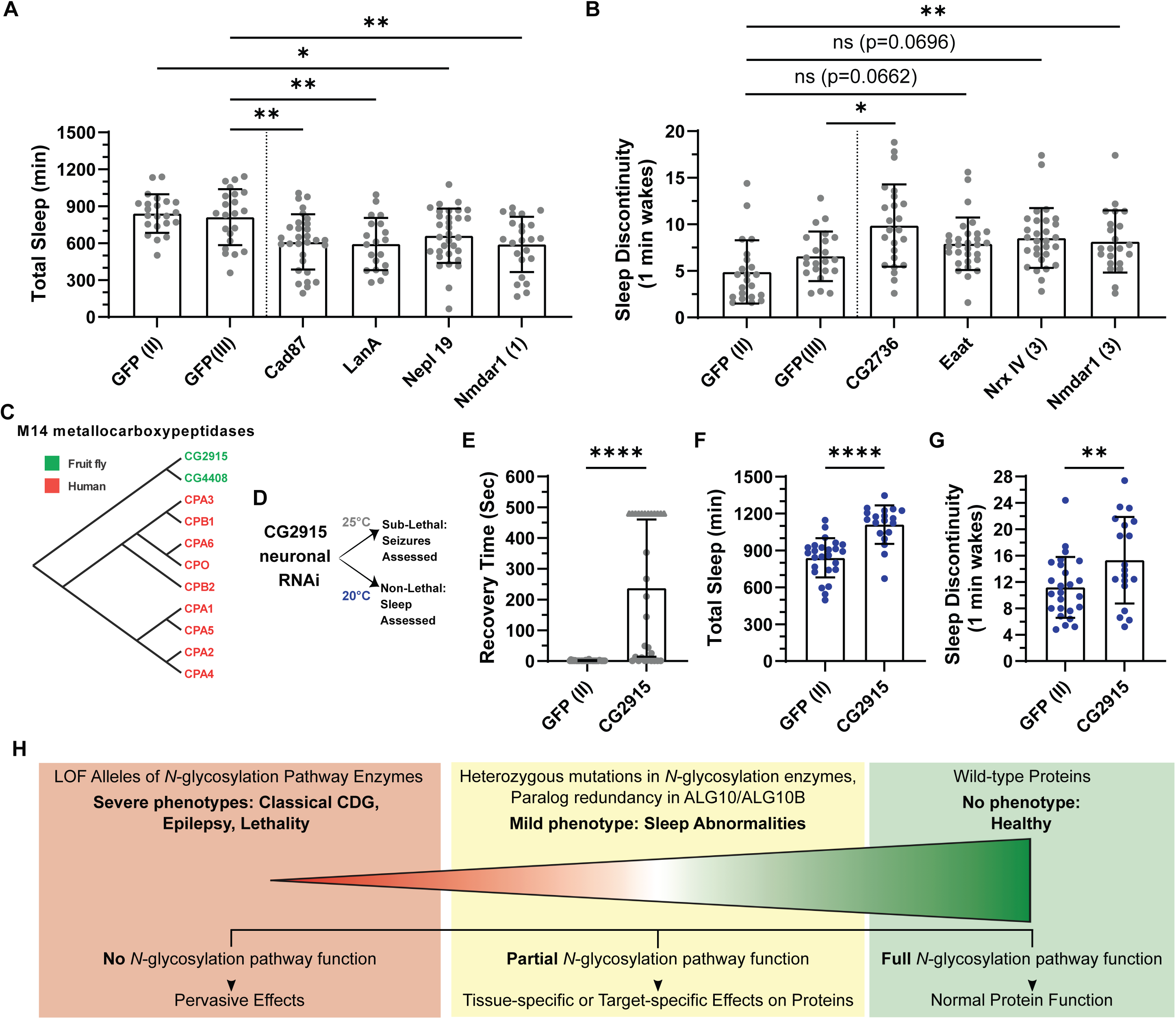
Functional assessment of the predicted targets of Alg10-mediated *N*- glycosylation in sleep and seizures. (A-B) Sleep duration and sleep discontinuity in control or neuronally restricted, gene-specific-knockdown of the indicated candidate targets of Alg10 glycosylation. Adjusted p-values from a one-way ANOVA with Šídák’s multiple comparisons test are shown. (C) Phylogenetic orthology of *dCPA1* (*CG2915*), a gene of unknown function in *Drosophila* showing a single paralog *CG4408* (based on Ensembl) and nine human orthologous M14 metallocarboxypeptidases. (D) Schematic detailing the temperature-based lethality of *dCPA1* neuronal knockdown and the phenotypic assessments carried out at each temperature. Sub-lethal refers to flies that are unable to survive the duration of the *Drosophila* locomotor activity and sleep monitoring assay. (E) Recovery time after bang-induced seizure in control or neuronal *dCPA1*-knockdown flies. Flies were observed for an upper limit of 8 minutes (indicated by a triangle data points). (F-G) Quantification of total sleep (F) and sleep discontinuity (G) in control and neuronal *dCPA1*-knockdown flies. Each dot in the graphs represents the mean for an individual fly over 5 days. Error bars represent the standard deviation. Blue dots indicate flies were raised at 20°C then assayed at 25°C in the locomotor activity and sleep monitoring assay. Adjusted p-values from a one-way ANOVA with Šídák’s multiple comparisons test are shown. (H) A model for the healthy, mild phenotypes, and severe phenotypes arising from respectively no, mild, or severe disruption of the *N*-glycosylation pathway.

Taken together, these experiments support the notions that reduced protein amount is, at least partly, responsible for the observed phenotypes when *N*-glycosylation of the protein is reduced. Furthermore, whereas knockdown of multiple target proteins led to sleep defects, knockdown of *dCPA1* (*CG2915*) was sufficient to recapitulate the sleep and seizure phenotypes present upon *Alg10* knockdown. These data suggest that although the reduced function of multiple target proteins may contribute to the sleep phenotypes, the dysregulation of dCPA1 may be a key regulator of both epilepsy-like phenotypes and sleep phenotypes.

## DISCUSSION

Our investigation of a human genetic locus for sleep and cardiovascular disease phenotypes spanning the two arms in the centromere-proximal regions of chromosome 12 led us to uncover a pair of paralogous *N*-glycosylation enzymes (ALG10 and ALG10B) that are required for neuronal homeostasis. *Drosophila* modeling supported a role for these enzymes in sleep regulation and uncovered a seizure phenotype. Driven by our phenotypic findings in flies, we subsequently identified an individual with progressive myoclonic epilepsy having an ultra-rare genotype of homozygous pathogenic/likely pathogenic variants in both *ALG10* and *ALG10B*. Underlying the conservation of *N*-glycosylation as a sleep regulator, disruption of *Alg10* orthologs in zebrafish and *C. elegans*, confirmed the presence of disrupted sleep observed in humans and flies. Additionally, a salt growth phenotype caused by disruption of the *Alg10* ortholog in *Arabidopsis thaliana* can be rescued by the overexpression of human *ALG10* indicating that the biochemical function of the ALG10 enzymes is conserved to Plantae. In further support of the role of *N*-glycosylation in these phenotypes, we provide evidence that multiple other enzymes in the *N*-glycosylation linear ‘assembly line’ pathway are involved in sleep and seizures in both humans and flies. Combining an unbiased proteomic and glycoproteomic strategy to uncover the proteins in the fruit fly brain that likely mediate the neurological symptoms, we provide evidence for their role in humans, and rigorously test the assumption that altered *N*-glycosylation alters protein abundance. In sum, our work highlights how experimental studies of genes from a GWAS locus in model organisms can not only support, but also guide characterization of novel human-disease relationships, and here, reveal mechanisms pointing to the significance of *N*-glycosylation in neurological and cardiovascular disease.

### Pericentromeric region of chromosome 12 displays common variant associations with sleep and cardiac traits

Human genetics has been an important tool for discovering genes for human sleep, particularly for rare variants ^47^. This report focuses on common variation with relevance to a larger fraction of the population and is one of the first where the underlying gene and mechanisms have been described in detail. Associations at the duplicated pericentromeric region harboring *ALG10/ALG10B* was consistently found across multiple sleep traits including self-reported morningness-eveningness preference, accelerometer-measured timing of the least-active 5 hours (L5) and timing of the most-active 10 hours (M10) of the day, self-reported sleep duration, and daytime napping, and was pleiotropic largely only to heart rate phenotypes, as a lead signal for resting heart rate as well as heart rate response to exercise ^48–50^. Notably, another common variant gene-mapping and cross-species mechanistic sleep study highlights a critical role for phosphatidylinositol glycan PIG-Q and glycosylphosphatidylinositol (GPI)-anchor biosynthesis in diverse neurons in sleep ^51^. Furthermore, *N*-glycosylation has been implicated in heart complications in 20% of CDGs ^52^, and remodeling of the cardiac myocyte glycome during development is key for regulation of cardiac electrical signaling^53^.

### *Drosophila* modeling reveals *N*-glycosylation as a regulator of the sleep-epilepsy axis

It has long been known that there is a link between sleep and epilepsy. For example, sleep deprivation is known to precipitate seizures in at-risk subjects, most of the deaths in SUDEP (<u>Su</u>dden <u>d</u>eath in <u>ep</u>ilepsy) occur during sleep ^54^, seizures in some forms of epilepsies are restricted to sleep or even specific sleep stages ^55^, specific sleep abnormalities are characteristic in epilepsies ^56^, and sleep disturbances (insomnia and excess daytime sleepiness) are common in epilepsy patients ^57–59^. A substantial body of work in model organisms such as *Drosophila* ^60–62^, zebrafish ^63^, and mice ^64–68^ recapitulates multiple aspects of these human observations. Despite the important implications of this sleep-epilepsy axis for human brain health, its molecular mechanisms have received limited investigation. Here, we uncover the *N*-glycosylation pathway as an important player on this sleep-epilepsy axis.

Upon modeling of the GWAS locus on chromosome 12, we found that knockdown of *Alg10* in *Drosophila* neurons revealed a conserved role in sleep behavior with significant increases in sleep and increased sleep discontinuity. Further, we discovered that neuronal *Alg10* knockdown revealed a propensity for bang-sensitive seizures with some animals also exhibiting spontaneous seizures. This bang-sensitive seizure phenotype has previously been shown in flies with mutations in voltage-gated potassium and sodium channels (e.g. *eag*, *sei*, *Sh*, and *para*) ^69^. Notably, the human orthologs of these genes (KCNH1, KCNH2, KCNA3, and SCN2A) are known to be glycoproteins, lending further support to the role of the *N*-glycosylation pathway in regulating the sleep-epilepsy axis. Knockdown of *Alg10* in subsets of neurons or specific brain regions using more restrictive Gal4 drivers found multiple drivers targeting the mushroom body and pars intercerebralis resulted in increased sleep. Increased sleep with bang-sensitive seizures were recapitulated with one cholinergic driver and one mushroom body driver indicating the disruption of the mushroom body may underly the *Alg10* knockdown sleep and seizure phenotypes.

### *Alg10 Drosophila* modeling helps identify a patient with ALG10/ALG10B deficiency

Currently, confirmation of a suspected CDG diagnosis in a patient is through sequencing of a panel of glycosylation pathway genes to detect coding variants. Despite sequencing thousands of patients, which has revealed a CDG associated with almost every gene in the *N*-glycosylation pathway, no disorder has been associated with the gene alpha-1,2-glucosyltransferase in humans. The duplication of the pericentromeric *ALG10/ALG10B* locus we describe in this work, which arose in an ancestor of our primate sublineage, may provide reasoning for the lack of an *ALG10*-associated CDG. Therefore, we hypothesized that loss of both ALG10 and ALG10B may be required for CDG-like symptoms to manifest, if these enzymes are functionally redundant.

Discovery of the seizure phenotype in *Drosophila* neuronal *Alg10* knockdown led us to explore the possibility of individuals with seizures having an increased chance of harboring variants in both *ALG10* and *ALG10B*. Within a cohort of patients diagnosed with progressive myoclonic epilepsy, we uncovered a report of an individual with a homozygous frameshift mutation in *ALG10* ^29^. Given the epileptic phenotype in this individual, our *Drosophila* modeling accurately predicted the presence of a variant also in *ALG10B* in this patient ^30^. Further, based on the similar neuronal phenotype present in the patient and *Drosophila* models, we predicted the patient might also have an undiagnosed sleep defect. This prediction was supported by polysomnography (PSG) assessment demonstrating the patient had difficulty maintaining sleep, while actigraphy data showed an increase in sleep latency and lower sleep efficiency of the patient compared to her family members. Our *Drosophila* modeling also provided an opportunity to functionally characterize the patient’s mutations in the context of sleep and seizure activity. While transgenic expression of either human *ALG10* or *ALG10B* rescues the total sleep and seizure phenotypes of the *Drosophila* neuronal *Alg10* knockdown, expression of the *ALG10B^L^*^253^*^W^* variant was able to fully rescue the bang-sensitive seizure phenotype and significantly rescue the total sleep, although not as well was either wild type *ALG10* or *ALG10B.* Contrastingly, transgenic expression of human *ALG10^K^*^391f^*^s^* failed to rescue either total sleep and bang-sensitive seizure phenotypes, while also worsening the sleep discontinuity phenotype. Together these findings suggest in the context of sleep and seizure regulation, *ALG10^K^*^391f^*^s^* is likely a null mutation and *ALG10B^L^*^253^*^W^*is likely a hypomorphic mutation. Not only did the *Drosophila* modeling allow for the sleep and seizure regulatory characterization of the patient’s variants, but we were only able to discover this individual due to our modeling work performed in *Drosophila*.

### Glycoproteomic analysis identifies brain relevant *N*-glycosylated targets that contribute to sleep and seizures phenotypes

Given the role of *N*-glycosylation as a post-translational modification, we aimed to identify the brain-relevant targets being affected in the neuronal *Alg10* knockdown flies that may be driving these sleep and seizure phenotypes. Our glycoproteomic profiling approach in *Drosophila* has two important conceptual distinctions from past catalogs of *N*-glycosylated proteins. Firstly, we used both wild-type and RNAi knockdown of a single *N*-glycosylation pathway enzyme in neurons, which enabled comparisons and improved the likelihood of identifying bona fide targets of the *N*-glycosylation pathway. Secondly, simultaneous *N*-glycoproteome and whole-proteome profiling has enabled us to test, at an omic scale, the dogma that reduced *N*-glycosylation leads to unfolding and destabilization of those target proteins. Most proteins with lower expression exhibited reduced *N*-glycosylation in *Drosophila* neurons, consistent with the directionality of our genetic knockdown (i.e., brain-specific RNAi of *Alg10*). Our primary motivation for the glycoproteomics was to identify and catalog *N*-glycosylated proteins in the brain that would be relevant to human CDGs. With this view, we focused on the targets whose role in human CDGs was supported by orthogonal human genetic or sequence conservation data. This led us to uncover multiple new proteins involved in sleep and epilepsy in humans and flies. Several of these are potentially actionable for therapeutics and will be the focus of future work. Furthermore, in performing our glycoproteomics analysis, to the best of our knowledge, we provide the first list of brain proteins in the fruit fly and conserved in humans whose *N*-glycosylation is critical for their normal function.

One target, *CG2915* (*dCPA1*), a member of the M14 family of zinc carboxypeptidase was found to play a vital role in *Drosophila* neuronal homeostasis, with RNAi knockdown resulting in a strong seizure phenotype. We note that mutations in human carboxypeptidase A6 are associated with juvenile myoclonic and generalized epilepsy ^70^. Additionally, RNAi knockdown of *dCPA1* resulted in an increase in sleep and increased sleep discontinuity similar to neuronal knockdown of *Alg10*. Two other targets, the glutamate receptor *nmdar1* and a predicted class B scavenger receptor *CG2736*, both resulted in increased sleep discontinuity upon independent neuronal RNAi knockdown. Furthermore, mutations in the human glutamate receptor, *GRIN1* are associated with neurodevelopmental disorder with hyperkinetic movements and seizures, and developmental and epileptic encephalopathy ^71,72^, while mutations in *SCARB2,* a class B scavenger receptor, are associated with progressive myoclonic epilepsy-4 ^73^ (**Table S25**). While these targets play essential roles in neuronal function, we assume it unlikely that a single protein is responsible for the sleep or seizure phenotypes observed upon *Alg10* knockdown.

### Modeling in other model organisms demonstrates a conserved sleep regulatory role and conserved enzymatic function of ALG10/ALG10B

The striking similarities between the impact of disrupted *N*-glycosylation in humans and *Drosophila* lead us to investigate the functional conservation of *Alg10* in other model organisms. Modeling of *alg10* loss in zebrafish revealed a decrease in nighttime sleep, characterized by shorter sleep bout lengths, while modeling of *algn-10* loss in *C. elegans* lead to an increase in sleep and an increase in the sleep-like state of lethargus. Notably, testing in *Drosophila* identified knockdown of some of *N*-glycosylated targets resulted in decreased sleep. It remains possible that the distinct effects in sleep phenotypes across our models may stem from differences in *N*-glycosylated targets between arthropods and vertebrates or targets may differ in their overall contribution to the sleep phenotype in different species. Although the directionality of the sleep disruption in *Alg10* knockdown in flies and *algn-10* nematodes differs with zebrafish *alg10* mutants, our modeling demonstrates a conserved function of *N*-glycosylation in general sleep regulation.

We were further interested in understanding the evolutionary conservation of *Alg10* beyond its role in sleep regulation. *N*-glycosylation has been previously associated with a leaf size phenotype and germination under salt stress in *Arabidopsis thaliana* ^39–42^. To test the enzymatic conservation of *Alg10*, we attempted to rescue the germination under salt stress phenotype of *alg10-1* mutant plants. Our work shows that not only does transgenic expression of either human ALG10 or ALG10B rescue the germination phenotype, but transgenic expression of ALG10B^L253W^ and ALG10^K391fs^ corroborate our characterization of the *ALG10* proband variants in *Drosophila* with *ALG10B^L^*^253^*^W^* rescuing the germination phenotype while *ALG10^K^*^391f^*^s^*fails to rescue.

## Conclusions

The broad impact of *N*-glycosylation on membrane and extracellular proteins underscores its importance in neuronal homeostasis. While this post-translational modification is a ubiquitous process found in all cell types, both our work here and that from others suggests that the heart and brain are the two major tissues affected by CDGs. Based on our glycoproteomic findings, we suspect that this may have to do with the preeminent role for *N*-glycosylation in controlling the function of integral membrane proteins involved in cellular excitability. It is noteworthy that cardiac and neuronal tissues are the two excitable tissue types in the metazoan.

Overall, this work integrating genetics, evolutionary history, and biochemical mechanisms at a human GWAS locus, is a case study in how *Drosophila* modeling can not only support, but also guide human-disease characterization, which points to the significance of *N*-glycosylation in neurological and cardiovascular disease. Moreover, it provides a molecular view of the previously recognized overlap of sleep disorders and epilepsy, details a roadmap for targeted therapeutic approaches for CDGs by generating a catalog of disease-relevant glycoproteomic targets of *N*-glycosylation, and highlights the evolutionary conservation of the biochemical function of the ALG10 enzymes across animals and plants. Ultimately, our work underscores how human genetics can help discover new biology.

While our study provides many biological and disease-relevant insights, we note several limitations. We cannot currently evaluate the directionality/causal effect on *ALG10* and *ALG10B* expression on the traits due to the common variant SNPs nearby. In our animal modeling we noted differences in directionality on sleep disruption between *Alg10* knockdown in flies and *algn-10* nematodes and the zebrafish *alg10* mutants. This may represent a fundamental difference in the identity and role of *N*-glycosylated proteins in sleep behavior between arthropods and vertebrates or between distinct sleep patterns such as the diurnal sleep pattern of zebrafish and the crepuscular sleep pattern of *Drosophila*. While our glycoproteomic analysis revealed neuronal *N*-glycosylated proteins, our survey is likely limited to abundant proteins with glycosylation sites within tryptic peptides. For example, we may have expected ion channels such as *Sh*, *sei*, and *para* as potential candidate target proteins given the strong seizure phenotype observed; however, these proteins have only a few predicted *N*-glycosylation sites and thus may not be enriched using this approach. Our *Drosophila* modeling focused primarily on neuronal manifestations of *N*-glycosylation. However, we also describe phenotypes related to reduced *N-*glycosylation in fly cardiac tissue, consistent with the heart trait associations of the *ALG10*/*ALG10B* locus and that CDGs have cardiac complications ^52,74^. In addition, given that our glycoproteomic analyses have so far been restricted to neuronal manipulations, we note that some of the human orthologs of our glycosylated targets have OMIM associations with cardiac-related disorders. Further characterization of these targets may also give insights into their roles and *N*-glycosylation in cardiac dysfunction.

## Data Availability

Data available at Mendeley Data.
The mass spectrometry proteomics data have been deposited to the ProteomeXchange Consortium via the PRIDE partner repository with the dataset identifier PXD055485.

https://doi.org/10.17632/kmf7bnxysd.1

https://doi.org/10.17632/w6jbwzvjt7.1

https://doi.org/10.17632/kn7z4z4cw5.1

## SUPPLEMENTAL FIGURE LEGENDS

**Suppl. Figure 1.**

(A-D) Regional association plots for the ALG10/ALG10B locus in GWAS of (A and B) chronotype (n=449,734) and (C and D) snoring (152,302 cases and 256,015 controls) in the UK Biobank population. The y-axis at left shows the -log10 P value for each variant in the region and the x-axis shows the genomic position. Each variant is represented by a filled circle, with the listed lead variant colored purple, and nearby variants colored according to correlation (r2) with the regional lead variant in the 1KG EUR population. The lower panel shows genes within the displayed region and the blue line corresponds to the recombination rate (right hand y-axis)

(E) Intrachromosomal duplication of a single ancestral gene located in the pericentromeric region (the centromere is shown in red) of human chromosome 12 gave rise to two orthologous genes: *ALG10* and *ALG10B*. Our estimates for the boundaries of this duplication shown with blue shading indicate that no other protein-coding genes are located in the region. All coordinates are with respect to the hg38 genome reference. Protein-coding genes are shown in blue, while non-protein-coding genes are in green.

(F) The karyotypic evolution of a single ancestral *ALG10* gene in the primate lineage. The underlying data are derived from Genomicus version 102.01 (**Table S2**). The vertical panel in light orange indicates the number of *ALG10*-like paralogs in the species. The speciation events in Hominoidea highlighted in red correspond to the duplication of an ancestral *ALG10*-like gene into two: *ALG10* and *ALG10B* around estimated 25 million years ago (Mya). Orthologs across species are indicated in a common color, while a light lilac color is used for other genes. The names for the human genes (and as a reference for the color scheme) are labeled on the genes in the row corresponding to humans. Gibbons (indicated with an *) have two *ALG10* paralogs but they are on different chromosomes. The species with a dagger sign next to their name are reversed with respect to their conventional chromosome orientations to align the *ALG10*-like gene along the human reference. Abbreviation: sp. = speciation event.

(G and H) Loss-of-function variants in *ALG10* and *ALG10B* are rare with no homozygotes. Allele frequencies (log10-transformed to aid visualization) of coding variants that are predicted to be loss-of-function in gnomAD for *ALG10* (top) and *ALG10B* (bottom) as a function of their location along the (G) genomic coordinate or (H) protein coordinate (normalized to 1) are shown. The highest frequency alleles (both frame shifts) are highlighted in red, to underscore their rarity in the population.

**Suppl. Figure 2. Characterization of *Drosophila Alg10* RNAi knockdown.**

(A) *Alg10* mRNA isolated from adult heads analyzed by RT-qPCR (n=20). The following genotypes were used: elav-Gal4 driving GFP^RNAi(III)^, Alg10^RNAi(W)^ and Alg10^RNAi(S)^. The relative normalized expression of *Alg10* was determined by comparing to *RpL32* before being expressed as fraction of expression in elav-Gal4>GFP^RNAi(III)^ control. Mean values from three technical replicates are shown with error bars showing the 95% confidence interval.

(B) Neuronal-specific expression of Alg10^RNAi(W)^ and Alg10^RNAi(S)^ using elav-Gal4 resulted in a reduction in life span compared to controls: elav-Gal4>GFP^RNAi(II)^ and elav-Gal4>GFP^RNAi(III)^. Graphs indicates the percent survival of adult males (n=>75 for each group) versus age post-eclosion (days).

(C) Cardiac-specific expression of Alg10^RNAi(W)^ and Alg10^RNAi(S)^ using Hand-Gal4, resulted in a significant reduction in lifespan compared to controls: Hand-Gal4/+ and Hand-Gal4>GFP-RNAi (p < .001). Graph indicates percent survival for adult females (n = 100 for each group) versus age post-eclosion (days).

**Suppl. Figure 3. Neuronal loss of Alg10 effect on circadian phenotypes and female sleep phenotypes.** (A-C) Quantification of circadian phenotypes in pan-neuronal and cry-Gal4 specific Alg10 knockdown using Alg10^RNAi(S)^. (D-F) Quantification of sleep parameters obtained from TriKinetics DAM system for individually housed flies. Each dot in the graphs represents the mean for an individual fly over 5 days. Error bars represent the standard deviation. The indicated parameter (y-axis) for strong (3rd chromosome) brain-specific knockdown of Alg10 in either virgin or mated female flies using an elav-Gal4 line are compared to the corresponding control (GFP-RNAi) transgenic strain. Adjusted p-values for the Šídák’s multiple comparisons test are shown.

Suppl. Figure 4. Characterization of the effects of Alg10 disruption on synaptic transmission and CPG properties at the 3^rd^ instar larval NMJ.

To explore if disruption of Alg10 function altered synaptic transmission, excitatory junctional potentials (EJPs) were recorded at *Drosophila* 3rd instar larval glutamatergic neuromuscular junctions (NMJs). (A) EJP amplitudes quantified from intracellular recordings at larval muscle 6. No significant differences were found between controls and RNAi knockdown of Alg10 (elav-GAL4>Alg10^RNAi(W)^ and elav-GAL4>Alg10^RNAi(S)^. Adjusted p-values from a one-way ANOVA with Dunnett’s multiple comparisons test are shown. Representative EJPs from each group are shown above: Blue, Oregon R; Green, Alg10^RNAi(W)^; Orange, Alg10^RNAi(S)^). Scale bar: 100 ms, 10 mV. (B) Mini frequency recorded from the indicated genotypes revealed no significant difference. Adjusted p-values from a one-way ANOVA with Dunnett’s multiple comparisons test are shown. Representative mEJPs from each group are shown above. Scale bar: 1.5 s, 4 mV. (C) Force-frequency plots generated by stimulating motor neurons at 11 different stimulation frequencies did not reveal significant differences across genotypes. Adjusted p-values from a one-way ANOVA with Dunnett’s multiple comparisons test are shown.

**Suppl. Figure 5. Variants in *ALG10* and *ALG10B***

Sequence chromatograms showing the location of the variants in *ALG10* and *ALG10B* in the proband (PME50) and the parents. The chromatogram of the *ALG10* variants is modified from Supplemental Figure S3 in ^30^.

**Suppl. Figure 6.** Germination features of *Arabidopsis alg10-1* mutant under different NaCl conditions with expression of *HsALG10* (A) Germination percentage (n=3-29) and (B) mean germination time (n=3-30) of *35s::AtALG10*, *35s::HsAlg10^WT^*, *35s::HsAlg10^K371fs^*, *35s::HsAlg10B^WT^*, and *35s::HsALG10B^L253W^* in the *alg10-1* mutant background under 0, 100, and 150 mM NaCl compared to Col-0 and *alg10-1* mutant. Data are mean ± SD, and n indicates the number of individual experiments. Adjusted p-values from a one-way ANOVA with Šídák’s multiple comparisons test are shown; ns = not significant at P 0.05, * = significant at P < 0.05, ** = significant at P < 0.01, *** = significant at P < 0.001, **** = significant at P < 0.0001.

Overexpression of *HsALG10* complements the seed germination phenotype of the *alg10-1* mutant under NaCl stress. Representative images of germinating seeds of *35s::AtALG10*, *35s::HsAlg10^WT^*, *35s::HsAlg10^K371fs^*, *35s::HsAlg10B^WT^*, and *35s::HsALG10B^L253W^* in the *alg10-1* mutant background under (C) 0, (D) 100, and (E) 150 mM NaCl over a seven-day period. 45 seeds of each genotype were plated on the agar media, and photos were taken from day 1 to day 7 after stratification. Number of germinating seeds is indicated at the right bottom of each box, and number of non-germinating seeds is in the bottom of each column.

**Suppl. Figure 7.** RT-qPCR confirming the overexpression of *AtALG10* and *HsALG10* in *Arabidopsis alg10-1* mutant. Relative normalized expression (ΔΔCq) of (A) *AtALG10* in Col-0, *alg10-1*, and *35s::AtALG10*, *35s::HsAlg10^WT^*, *35s::HsAlg10^K371fs^*, *35s::HsAlg10B^WT^*, and *35s::HsALG10B^L253W^* in the *alg10-1* mutant background, (B) *HsAlg10^WT^* in *35s::HsAlg10^WT^*in the *alg10-1* mutant*, (C) HsAlg10^K371fs^* in *35s::HsAlg10^K371fs^*in *alg10-1* mutant, *(D) HsAlg10B^WT^* in *35s::HsAlg10B^WT^*in *alg10-1* mutant, and (E) *HsALG10B^L253W^* in *35s::HsALG10B^L253W^*in *alg10-1* mutant. Data are mean ± SD, and n = 3-4 biological replicates. The expression was normalized by *ACTIN2* (*ACT2*), *ISOPENTENYL DIPHOSPHATE ISOMERASE 2* (*IPP2*), and ArrayControl RNA spikes (Invitrogen, USA). In (A), adjusted p-values from a one-way ANOVA with Šídák’s multiple comparisons test are shown; ns = not significant at P 0.05, * = significant at P < 0.05, ** = significant at P < 0.01, *** = significant at P < 0.001, **** = significant at P < 0.0001. Replicates of immunoblot analysis of *Arabidopsis* using anti-HRP to detect glycoprotein content (F and G). Immunoblot analysis of total protein crude extracts from Col-0, *alg10-1*, and *35::AtALG10*, *35::HsAlg10^WT^*, *35::HsAlg10^K371fs^*, *35::HsAlg10B^WT^*, and *35::HsALG10B^L253W^* in the *alg10-1* mutant background using anti-HRP. Proteins were separated on 7.5% SDS-PAGE, and the blot stained with Ponceau S solution prior to immunodetection. Biological replicates of blots shown in Figure 4P. Glycoprotein detection in *Arabidopsis* using lectin blots. (H-J) Lectin blots of total protein crude extracts from Col-0, *alg10-1*, *35s::AtALG10*, *35s::HsAlg10^WT^*, *35s::HsAlg10^K371fs^*, *35s::HsAlg10B^WT^*, and *35s::HsALG10B^L253W^*in the *alg10-1* mutant background using biotinylated concanavalin A which binds to mannose residues on glycoproteins. Proteins were separated on 7.5% SDS-PAGE, and the blot was stained with Ponceau S solution prior to incubation with Concanavalin A (Con A). The asterisks indicate the difference in protein bands between Col-0 and *alg10-1* mutant. Three biological replicates are shown.

**Suppl. Figure 8.**

(A-E) Regional association plots for the *SCL39A8* and *DOLPP1* loci in GWAS of short sleep duration (n=106,192 cases, 305,742 controls), sleep duration (n=446,118), excessive daytime sleepiness (n=452,071), naps (n=452,633) and chronotype (n=449,734) in the UK Biobank population. The y-axis at left shows the -log10 *P* value for each variant in the region and the x-axis shows the genomic position. Each variant is represented by a filled circle, with the listed lead variant colored purple, and nearby variants colored according to correlation (r^2^) with the regional lead variant in the 1KG EUR population. The lower panel shows genes within the displayed region and the blue line corresponds to the recombination rate (right hand y-axis).

## SUPPLEMENTAL TABLES

**Table S1:** Compilation of GWAS hits at or near human *ALG10* and *ALG10B* from published studies. The results are sorted by p-value for the genetic association (column E). Results from an analysis by Ben Neale are cited as a URL (column H), published results are cited as a PMID while unpublished or publications from our labs that are in review are cited as ‘unpublished’ or ‘interim’. neg_log_p_value (column F) is the negative logarithm in base 10 of the p-value in column E. Standard nomenclature is used for other column titles.

**Table S2:** Genomicus results for human *ALG10* and *ALG10B*. The number of species (column B), estimated timeframe (column C, Mya = million years ago) and the number of homologs of the gene in question (column D) are as shown for the lineage in the evolutionary tree corresponding to *Homo sapiens* (rows 26 and 58), starting with metazoans (rows 2 and 34). AlignView refers to an alignment between gene orders in different species. PhyloView refers to the gene order of a reference gene and its neighbouring genes, and the order of their respective orthologs and paralogs in different species that share the same ancestral “root” species. For more details, please see Genomicus’ help and documentation (the Views tab) at https://www.genomicus.bio.ens.psl.eu/

**Table S3:** Orthologs of *ALG10* and *ALG10* in multiple primate species

A list of extant primate species (first column) and the ENSEMBL identifiers for the ALG10-like genes in that species are provided.

**Table S4:** Seizure and sleep phenotyping of Alg10^RNAi^ ^(W)^ and Alg10^RNAi^ ^(S)^ along with their corresponding GFP RNAi controls. Seizure recovery time was measured manually. Sleep parameters were measured using ClockLab and a custom R script. The data are visualized in Figure 2.

**Table S5:** MARGO-based locomotion phenotyping of Alg10^RNAi^ ^(W)^ and Alg10^RNAi^ ^(S)^ along with their corresponding GFP RNAi controls. Raw speed data is in pixels change per frame and speed data is in millimeters per sec. The data are visualized in Figure 2G.

**Table S6:** Electrophysiological phenotyping of Alg10^RNAi^ ^(W)^ and Alg10^RNAi^ ^(S)^ along with Ore^R^ as a control. Electrophysiological readings were collected from third-instar larvae. The data are visualized in Figure 2I and 2J.

**Table S7:** Sleep and seizure phenotyping of brain region- and neuronal subset-specific knockdown of Alg10 using different brain Gal4 drivers. Seizure phenotypes were assessed as present or not via a quick bang of the vial on the working surface. Sleep data is shown as both normalized and raw total sleep values. The data are visualized in Figure 2K.

**Tables S8 and S9:** Predicted loss-of-function alleles of ALG10 (Table S8) and ALG10B (Table S9), their annotations and descriptors, as well as data describing their frequency in the healthy gnomAD cohort are as shown.

**Table S10** Summary of actigraphy results from proband and immediate family member

**Table S11:** Locomotor activity and sleep in zebrafish *alg10* homozygous mutant, heterozygous mutant, and wildtype sibling controls during the day and night. The data are visualized in Figure 4.

**Table S12:** Lethargus duration and total sleep data for *C. elegans algn-10*(*ok809);* this gene is also known as *tag-179* or *T24D1.4*. The data are visualized in Figure 4.

**Table S13:** Germination information for *HsAlg10* in Arabidopsis

Germination index of Col-0, *alg10-1*, and *35s::AtAlg10*, *35s::HsAlg10^WT^*, *35s::HsAlg10^K371fs^*, *35s::HsAlg10B^WT^*, and *35s::HsAlg10B^L253W^* in the *alg10-1* mutant background. The data are visualized in Figure 4O. Mean Germination Time and Germination Percentage data for the same germplasm for germination index are visualized in Figure S6.

**Table S14:** Orthologs of human *N*-glycosylation pathway genes (columns A, B, C) in the fly *Drosophila melanogaster* (columns E and F) as obtained from DIOPT along with quality metrics (columns G to K) and source databases (column L).

**Table S15:** Genotypes and Bloomington Drosophila Stock Center numbers for RNAi lines targeting *N*-glycosylation pathway genes.

**Table S16**: Seizure and sleep phenotyping of neuronal knockdown of *N*-glycosylation pathway enzymes via RNAi along with their corresponding GFP RNAi controls. Seizure recovery time was measured manually. Sleep parameters were measured using ClockLab and a custom R script. The data are visualized in Figure 5B-D.

**Table S17:** Cardiac phenotyping of heart-specific knockdown of Alg10 and other *N*-glycosylation pathway enzymes, using Hand-Gal4, along with driver only control. The data are visualized in Figure 5F-H.

**Table S18:** P values of Pascal (Pathway Scoring Algorithm) for human *N*-glycosylation enzymes in nine sleep and chronotype traits.

**Table S19:** UKB ALG Pathway. *N*-glycosylation pathway genes on haplotypes associated with sleep and chronotype traits. The lead SNP is shown in column A while the genes in linkage disequilibrium with this lead SNP are in column D. The nearest gene to the lead SNP is indicated with an asterisk. *N*-glycosylation genes are in bold.

**Table S20:** Glycoproteomics - protein level. Spectral counts of detected *N*-glycosylated proteins combined over the entire protein in wild wttype (WT), Alg10 RNAi (S), or Alg10 (VS) *Drosophila* head samples.

**Table S21:** Glycoproteomics - site level. Spectral counts of individual *N*-glycosylation sites of proteins in wild type (WT), Alg10 RNAi (S), or Alg10 (VS) *Drosophila* head samples. The detected *N*-glycosylated protein fragment is in column B with the glycosylated asparagine annotated as a lower case “n”.

**Table S22:** Proteomics. Spectral counts of detected proteins in wild type (WT), Alg10 RNAi (S), or Alg10 (VS) *Drosophila* head samples.

**Table S23:** Glycoproteomics - prot lvl stats. The fold change of *N*-glycosylated proteins and the P values between Alg10 and WT samples.

**Table S24:** Glycoprot site lvl stats. The fold change of individual *N*-glycosylation sites and the P values between Alg10 and WT samples.

**Table S25:** Sleep and seizure phenotyping of neuronal knockdown of *N*-glycosylation targets identified by glycoproteomics as having altered expression levels upon neuronal- knockdown of Alg10. The data are visualized in Figure 7A-B and Fig 7E-G.

**Table S26:** Human orthologs of altered proteins from fly brain glycoproteomics ranked based on DIOPT score are listed. Genetic associations with monogenic epilepsy or epilepsy-like neurological traits in OMIM are shown in column G. A more exhaustive list of all monogenic disorders associated with each ortholog is shown in column J . GWAS results from the NIH GWAS catalog for epilepsy, sleep and all phenotypes are listed in columns H, I and K respectively.

## MATERIALS AND METHODS

### Data and Code Availability

Drosophila analysis code is available on GitHub at https://github.com/jameswalkerlab/Gill_et.al.

Zebrafish analysis code is available on GitHub at https://github.com/proberlab/videotracker-scripts

*C. elegans* sleep analysis software was previously described ^86^ and code was deposited at https://github.com/Huiyan-Huang

### Human Genetics

SNPs at or near *ALG10* and *ALG10B* with genome-wide significant associations with human traits were curated from PheGeni ^91^ and PhenoScanner ^92,93^ and manually categorized into sleep/circadian, cardiovascular, and other traits. SNP annotations were obtained from dbSNP ^94,95^ and openSNP using the R packages LDlink and rsnps ^96^. Locuszoom ^85^ was used to visualize GWAS results, and the PASCAL software tool ^84^ was used to perform gene-based analyses using summary statistics for sleep traits downloaded from the Sleep Disorder Knowledge Portal (sleep.hugemap.org).

Gene-based analysis of genes encoding N glycosylation enzymes was performed using PASCAL ^84^, a pathway scoring algorithm that computes gene-based scores based on GWAS summary statistics. The gene score and associated p-value was calculated for each sleep trait based on the average association signal per gene (sum option) summing independent signals per gene.

### Patient methods

All human studies on the epilepsy proband and relatives were performed with informed consent. We received ethics approval for the sleep studies.

#### Variant validation and segregation

Variants in *ALG10* and *ALG10B* were confirmed by bi-directional Sanger sequencing (ABI BigDye 3.1, Applied Biosystems) on ABI3730xl DNA Analyzer. Primers were designed with Primer-BLAST ^97^. The sequences were analyzed using Sequencher v.5.3 (Gene Codes Corporation). Primers for PCR and Sanger sequencing are available upon request.

#### Brain imaging

Brain imaging was performed via a 1.5 T MR machine, sagittal and axial T1, axial and coronal T2, axial FLAIR, diffusion and ADC sequences were acquired. The images were evaluated by neuroradiology specialists.

#### Polysomnography (PSG), actigraphy and R-R interval variability assessment

PSG recording was performed according to AASM (American Academy of Sleep Medicine) manual version 2.5 (current at the time of the PSG) for recording and scoring of sleep and associated events. Additional sixteen channels of EEG were involved in the PSG because of the primary diagnosis of epilepsy and to evaluate the micro-organization of sleep in detail. Two-channels of electrooculography, surface electromyography recordings from chin and bilateral tibialis anterior muscles, body position, electrocardiography (Lead II), snoring sensor, oro-nasal thermistor, nasal pressure, thoracal and abdominal belts for respiratory movements, pulse oximetry, and synchronous video monitoring were recorded.

Sleep and sleep-related events were scored by G.B.S. (Sleep Expert by the European Sleep Research Society) on the basis of AASM Manual (version 2.5), and hypnogram was automatically formed by the Grass® Technologies. Sleep disorders were diagnosed based on the 3^rd^ edition of the International Classification of Sleep Disorders (ICSD-3) published by AASM in 2014.

Sleep was objectively assessed using wrist actigraphy with the Philips Actiwatch Spectrum Pro (Philips Respironics), a validated tool worn on the nondominant wrist that measures wrist movement and ambient light. A standardized scoring approach, as described previously, was used to estimate sleep parameters ^98^. Briefly, rest intervals were ascertained based on light intensity and activity levels as sleep logs were not available. For each determined rest interval, the Actiware software (version 6.0.9) then determined sleep and wake status (i.e. sleep onset, offset, and wakefulness in bed) to calculate sleep onset latency and total sleep time. To estimate habitual sleep, we averaged estimates for all sleep measures across all days assessed for each participant.

R-R interval analysis was performed with the subject relaxed and in supine position. Recordings during rest and hyperventilation were acquired via an EMG system (Natus Keypoint v2.4). Disposable cup electrodes were positioned on the extremities to gather recordings. Filters were set to 5 Hz and 100 Hz. Normal values for the R-R interval variation according to age was derived from ^99^

#### Drosophila melanogaster

All *Drosophila* stocks used in this study are listed in the key resources table. Flies were raised on standard cornmeal medium with a 12h/12h light cycle at 25°C with a relative humidity 50-70%, unless otherwise indicated. Strains used in this study include: Oregon-R-P2 (Oregon R, Bloomington Drosophila Stock Center BDSC: 2376), elav-Gal4: P{GawB}elav[C155] (BDSC: 458) and Hand-Gal4^75^ (gift from Eric Olsen).

#### Drosophila locomotor activity and sleep monitoring

*Drosophila* sleep was monitored by locomotor activity by quantifying infrared beam breaks using the Drosophila Activity Monitoring (DAM) System (Trikinetics, Waltham MA). Individual 3-9-day old male flies were loaded into glass tubes containing standard fly food and monitored for 5 days at 25°C. Collected data was analyzed using ClockLab and RStudio. Sleep in *Drosophila* was defined as any period of behavioral quiescence five minutes or longer as previously described ^100,101^. Sleep Discontinuity was defined by the number of 1-minute wakes during a 24-hour period. Dusk activity was measured between ZT11 and ZT13. Dawn activity was measured between ZT23 and ZT1. At least 20 flies of each genotype were tested except for several genotypes which were sub-viable with few flies surviving the entire 5-day LD period (see **Tables S4, S16, S25)**. In these cases, as many flies as possible, that survived the 5-day LD period, were analyzed. If a genotype was not viable when raised at 25°C, flies were raised at 20°C. For female experiments, virgin flies of the corresponding phenotype were identified by the presence of a visible meconium. Female flies without a visible meconium were collected and placed in a vial for a day with male flies of the same genotype to ensure mating. Individual 2-4 day old virgin and mated females were loaded into the DAM system in the same manner as male flies described previously. However, female flies were monitored for 4 days to avoid erroneous beam breaks caused by larvae that may begin wandering out of the food beyond day 4. All sleep parameters were quantified for females as previously described for males. DAM system data was collected and analyzed in RStudio using a custom code. RStudio scripts and methodology can be found on https://github.com/jameswalkerlab/Gill_et.al. One-way ANOVA with Šídák’s multiple comparisons test for DAM system data and Bang-sensitive seizure data were performed using GraphPad Prism.

#### Drosophila MARGO locomotion monitoring

*Drosophila* locomotion was video monitored using the MARGO system for automated tracking (Werkhoven et al., 2019) and behavioral arena boards that allowed tracking over multiple days ^26^. Behavior boards were prepared by the addition of standard fly food at one end. Individual 3-7-day old male flies were loaded into the behavior boards such that there was one fly per arena. Fly behavior was monitored via a digital camera and fly centroids were tracked in real-time for 3 days at 20°C. Collected data was analyzed using a custom script in MATLAB. Speed was calculated as the change in position of the centroid of the tracked fly between frames. This data was converted to millimeters per sec using an experimentally derived conversion factor of 3.4667 pixels per millimeter. Since the video was acquired at at 4 Hz, this value was used to convert millimeters per frame data to millimeters per seconds. For average speed calculations, frames where flies were immobile were excluded. Therefore, average speed measurements are representative of the average speed while flies are in motion.

#### Drosophila circadian rhythm monitoring

*Drosophila* circadian rhythm was monitored using the DAM system under constant darkness. Individual 3-9-day old male flies were loaded into glass tubes containing standard fly food. Flies were entrained for 4 days under 12:12 LD conditions at 25°C then transitioned to constant darkness for 5 days at 25°C. Locomotor activity from days 1 through 5 of constant darkness were analyzed for circadian phenotypes. Collected data was analyzed using ClockLab and RStudio. Rhythmicity was scored as previously described ^102^ with few modifications. Briefly, the first criteria for rhythmicity versus arrhythmicity was a rhythm with 95% confidence using Χ^2^ analysis of the periodogram. The second criteria stratified the strength of rhythmicity using the corresponding fast Fourier transform value (FFT). The rhythm stratification parameters we categorized as strong (FFT > 0.855), moderate (FFT = 0.0061-0.588), weak (FFT = 0.0042-0.006), and arrhythmic (FFT < 0.0042). The average FFT and average period were calculated for rhythmic flies only.

#### Drosophila Bang-sensitive paralysis behavioral testing

Behavioral testing for bang-sensitive (BS) paralysis was performed as previously described ^103^ with minor modifications. Flies 1-6 days post-eclosion were collected under CO_2_ and placed in a clean vial, with up to 25 flies in a single vial. Flies were allowed to recover for at least 1 hour after transfer. Flies were then vortexed on a Talboys standard multi-tube vortexer (945007) at a vortex speed of 7 for 10 seconds before filming with an IPhone. Videos were analyzed using QuickTime Player (Apple). For each fly, the recovery time from BS paralysis was determined by recording the amount of time it took a fly to stand after vortexing. Flies that did not recover after 8 minutes were recorded at a maximum recovery time of 480 seconds but annotated as having not recovered. If a genotype was not viable when raised at 25°C, flies were raised at 20°C and tested for BS paralysis or annotated as lethal. For the analysis of BS paralysis phenotypes of elav >Alg10^RNAi^ knockdowns, at least 50 flies were tested; for analysis of BS paralysis phenotypes in the knockdown of other *Alg* enzymes at least 20 flies were tested except for *Alg2* RNAi (BDSC: 55671) and *xit* RNAi (BDSC: 55190), which were sub-viable and therefore as many flies as possible that survived 1-day post-eclosion were tested. For the analysis of RNAi knockdowns of the targets of N-glycosylation, an initial screen for evidence of bang-sensitive seizures was conducted; follow up assays with quantification for those genotypes with seizures were performed with at least 20 flies.

#### Drosophila Reverse Transcription Quantitative PCR (RT-qPCR)

For each genotype, RNA was extracted from 20 adult fly heads (equal mix of females and males) using TRIzol Reagent (Invitrogen). Pellets were resuspended in RNase-free water and treated with the DNA-free DNA Removal Kit before cDNA was produced using the ProtoScript II kit (NEB). qPCRs were run using FastStart Universal SYBR Green Master (Rox) and LightCycler 480 (Roche). Relative normalized expression (ΔΔCq) of *Alg10* was determined comparing to *RpL32*. Three technical replicates were performed for each cDNA sample for each target gene.

#### Drosophila Electrophysiology

Wandering 3^rd^ instar larvae were isolated from the sides of culture vials and dissected in modified hemolymph-like HL3 saline containing (in mM): NaCl: 70; KCl: 5; CaCl_2_: 0.25; MgCl_2_: 20; NaHCO_3_: 10; Trehalose: 5; Sucrose: 115; HEPES: 5, pH = 7.18 ^104^. Larvae were dissected to access dorsal muscle fibers and nerves were severed as they emerged from the ventral nerve cord. Excitatory junctional potentials (EJPs) were elicited by stimulating severed abdominal nerves. A Master 8 A.M.P.I. (Jerusalem, Israel) stimulator was used to elicit stimulation via a suction electrode (A-M systems, Sequim, WA). EJPs were recorded using sharp glass micro-electrodes containing a 2:1 mixture of 3M potassium chloride: 3 M potassium acetate, with electrode resistances of 40-80 MΩ. An Axoclamp 2B amplifier (Molecular Devices, San Jose, CA) was used for signal detection and data was digitized via Axon Instruments Digidata 1550 (Molecular Devices). Signals were acquired at 10 kHz using Clampex and processed using Clampfit and MiniAnalysis. Temperature was controlled using a CL-200A heater-cooler controller (Warner Instruments, Hamden CT) with a heat sink (Koolance) by constant perfusion of physiological saline through a Harvard apparatus in-line saline temperature controller (Holliston, MA) and a Warner Instruments dish temperature regulator (Model TB3 CCD).

#### Nerve-evoked contraction force recordings

All force recordings were obtained using an Aurora Scientific 403A force transducer system (Aurora Scientific, Aurora, Canada), which includes a force transducer headstage, an amplifier, and digitizer. Nerve-evoked contractions were generated using bursts of electrical stimuli from a Master 8 (A.M.P.I.) stimulator. Saline composition was similar to the electrophysiological recordings but with the following changes: CaCl_2_: 1.5mM; MgCl_2_: 4mM. The duration of single impulses was 5x10^-4^ s and the interburst duration was kept constant at 15 s. Burst duration and frequency were altered for each individual experiment as previously described ^105^. Digitized data was acquired using Dynamic Muscle Acquisition Software (DMCv5.5). Digitized data was imported and processed in MATLAB using custom code.

#### Drosophila Cardiac imaging

Flies of both sexes were imaged at 3 weeks post-eclosion. In these semi-intact heart preparations, nerve input is eliminated so that endogenous cardiac pacing can be observed. Direct immersion optics was used in conjunction with a digital high-speed camera (up to 150 frame/s) to record contraction movements using HCImage software. Fly hearts were bathed in artificial hemolymph and imaged by a high-speed imaging system. A SOHA-based image analysis software ^106,107^ was used to calculate cardiac function parameters such as heart rate (HR), systolic diameter (SD), systolic interval (SI), diastolic diameter (DD), diastolic interval (DI), arrhythmia index (AI), and fractional shortening (FS). These parameters enable interpretations on the frequency of heartbeat per second (HR); the diameter of the beating heart at full relaxation (DD) and contraction (SI); the time length of relaxing status (DI) and contracting status (SI); variability of heartbeat length (AI) and contractility (FS). For the determination of cardiac parameters, at least 15 hearts were analyzed. SOHA-based image analysis software was used as previously described ^106,107^ to quantify HP, DD, SD, DI, SI, AL and FS. One-way ANOVA with Dunnett’s multiple comparisons test were performed using GraphPad Prism.

#### Drosophila transgene construction

Reference sequences from Gateway donor clones for *ALG10* cDNA clone (IOH21803 corresponding to Ref seq. NM_032834.3) and *ALG10B* cDNA clone (IOH55327 corresponding to Ref seq. NM_001013620.3) in pDONR221 vector were transferred to pGW-HA-attB using standard Invitrogen Gateway LR cloning protocol (11791020). To generate UAS-variant cDNAs the constructs are mutagenized in donor vectors using NEB Q5 Site-Directed Mutagenesis Kit (E0544S) using the manufacturer’s protocol. Site directed mutagenesis using primers Alg10-K371fs-FOR/Alg10-K371fs-REV and Alg10B-L253W-FOR/Alg10B-L253W-REV was used to generate the Alg10^K371fs^ and *ALG10B^L^*^253^*^W^* variants respectively. Variant cDNAs are sequenced and transferred to pGW-HA-attB vector using standard Invitrogen Gateway LR cloning protocol (Cat #: 11791020). The wild-type and mutant cDNAs are inserted in the VK37 landing site (PBac{y^+^-attP-3B}VK00037) using phiC31 mediated site specific transgenesis.

#### Generation of Alg10 RNAi line from dna2417

Since no Alg10 RNAi lines at the Vienna Drosophila Research Center (VDRC) were available, we obtained the pMF3 UAS-Alg10 RNAi plasmid (dna2417) from the VDRC GD RNAi library and used this for P-element-mediated germline transformation by injection into *w*^1118^ embryos (Rainbow Transgenic Flies, Inc. Camarillo CA). A transgenic line was established, mapped to the third chromosome, and balanced with TM3 using standard protocols.

#### C. elegans

*C. elegans* strains used were N2 (Caenorhabditis Genetics Center) as a wild type control and VC569, algn-10 (ok809)/hT2 bli-4(e937) let-?(q782) qIs48 (C. elegans Gene Knockout Program)^77,78^. *C. elegans* were raised at 20°C on NGM agar plates and fed *E. coli OP50* ^88^. Animals in the L4.2 to L4.5 substages were selected for experiments ^89^. The VC569 strain (*algn-10 (ok809)/hT2 bli-4(e937) let-?(q782) qIs48*) was created by the International *C. elegans* Gene Knockout Consortium (doi:10.1534/g3.112.003830) but the deletion location has not described previously. Sequencing using primers listed in table were used to determine the allele is a 630bp deletion that removes part of exon 4, intron 4 and more than half of exon 5. The deletion is externally flanked by sequences TACCGATTTGTTGTCACTCACTTCAGTTAT and TATTCCACAAAATTCTATTTATAATCGCAT.

#### C. elegans sleep assessment

A previously described ^86^ microfluidic system was used to evaluate *C. elegans* developmentally timed sleep during the transition between the L4 stage and adulthood. An *E. coli OP50* culture was treated for at least a week with kanamycin at 4°C, then resuspended in liquid NGM (final OD_600_ of 6.7). This bacterial culture was added to microfluidic chip chambers as a food source. Mid-L4 stage animals were gently picked into individual chambers. Chambers were covered with a glass coverslip and sealed using 2% agar. Images were captured every 10 seconds for 12 hours.

Lethargus duration and total sleep were calculated using a MATLAB image subtraction script ^38^ and a custom Python script ^86^. Entry into lethargus was defined as the point at which the fractional quiescence, calculated using a rolling average over 10 minutes, stayed above 0.1 for at least 20 minutes. Exit from lethargus was defined as the point at which fractional quiescence stayed below 0.1 for at least 20 minutes. Total sleep was defined as the sum of time an animal spent in quiescent bouts (with a duration of at least 10 seconds) during lethargus. Lethargus duration was defined as the time elapsed between entry to and exit from lethargus. One-way ANOVA with Dunnett’s multiple comparisons test were performed using GraphPad Prism.

#### Zebrafish

Zebrafish lines used were AB (Zebrafish International Resource Center (ZIRC) Catalog ID: ZL1) and TL (ZIRC Catalog ID: ZL86. The *alg10 d16* mutant line (*alg10+/-*) contains a 16 base-pair deletion, resulting in a shift of the translational reading frame after amino acid 71, which is predicted to produce a 77 amino acid truncated mutant protein compared to the 473 amino acid wild-type protein. The predicted mutant protein lacks transmembrane domains 2 - 12 and is thus likely to be non-functional. The TLAB wild-type strain was generated by mating fish of the TL and AB strains, which were obtained from the Zebrafish International Resource Center (ZIRC). All experiments involving zebrafish were performed in accordance with the Institutional Animal Care and Use Committee (IACUC) guidelines and by the Office of Laboratory Animal Resources at the California Institute of Technology (animal protocol 1836).

#### Generation of zebrafish alg10 mutant

A TBLASTN search of the human ALG10B amino acid sequence against the zebrafish genome identified a single *alg10* ortholog for human ALG10 and ALG10B. Using CRISPR/Cas9, the zebrafish *alg10* mutant was generated as previously described ^108^. Cas9 protein (Integrated DNA Technologies) and a sgRNA (5’-TGGGCTGTATTTGGCATCGG-3’) targeting the second transmembrane domain in the second exon of the *alg10* gene were co-injected into zebrafish embryos at the one-cell stage. Injected animals were raised to adulthood and animals carrying germline mutations were identified by PCR fragment analysis ^109^. Mutant founders were outcrossed to the WT parental TLAB strain and their progeny were genotyped by PCR to identify mutant animals. Sanger sequencing was then used to determine the exact nature of the mutation. Mutants were genotyped using the zebrafish *alg10* forward primer and zebrafish *alg10* reverse primer which produce a 188 bp PCR product for the wild-type allele and a 172 bp PCR product for the mutant *alg10* allele. *alg10* heterozygous mutants were outcrossed to the parental TLAB wild-type strain for 2 generations prior to behavioral experiments to remove potential off-target mutations. Each experiment was performed blind to genotype and animals were genotyped by PCR after each experiment was complete.

#### Zebrafish behavioral assays

Sleep and locomotor activity assays were performed as previously described ^34^. *alg10 +/-* adult zebrafish were in-crossed to generate *alg10 +/+, +/-, and -/-* siblings, which were raised on a 14:10 hour light:dark cycle in E3 medium (5 mM NaCl, 0.17 mM KCl, 0.33 mM CaCl_2_, 0.33 mM MgSO_4_, pH 7.4). Starting at 4 days post-fertilization, individual larvae were placed in each well of a 96 well plate blind to genotype and filled with 650 µL of E3 medium. The plate was sealed with optical adhesive film (4311971, Applied Biosystems) to prevent evaporation. The sealing process sometimes introduces air bubbles in some wells, which were discarded from analysis. Behavior was monitored using a customized Zebrabox (Viewpoint Life Sciences) and an automated video tracking system with a Dinion one-third inch monochrome camera (Dragonfly 2, Point Grey) fitted with a fixed-angle megapixel lens (M5018-MP, Computar) and infrared filter. The 96-well plate was illuminated from below using white LEDs from 9 am to 11 pm and continuously illuminated with infrared LEDs. The plate was placed inside a chamber with recirculating water to maintain a temperature of 28.5°C. Behavior was recorded at 15 Hz using quantization mode in 1-minute time bins using the following empirically determined parameters: detection threshold: 15; burst: 29; freezing: 3. A movement bout was defined as a pixel change between adjacent video frames preceded and followed by a period of inactivity of at least 67 ms (the limit of temporal resolution). Sleep was defined as one or more minutes of inactivity because this is associated with a significant increase in arousal threshold ^34^. A sleep bout was defined as a continuous string of sleep minutes.

Data were processed using custom MATLAB scripts and statistical tests were performed using Prism (GraphPad). One-way ANOVA with Dunnett’s multiple comparisons test were performed using GraphPad Prism.

### Arabidopsis

Arabidopsis Columbia (Col-0) accession was used as a wild-type. *alg10-1* (SAIL_515_F10) seeds were ordered from ABRC (Arabidopsis Resource Center) ^90^. PCR was performed to genotype *alg10-1* (using SAIL_515_F10_F, SAIL_515_F10_R, and LB3), and homozygous plants were selected for propagation. Seeds were sterilized by 50% (v/v) bleach and 0.01% (v/v) Triton-X and sown on ½ Murashige and Skoog (MS) (Phytotech Labs, USA) supplemented with 0.4% (w/v) agar. Seeds on plates were stratified at 4°C for three days and subsequently transferred to the growth chamber with 12/12 hours light/dark cycles and constant 23°C. For seed propagation, two- or three-week-old seedlings were transplanted to soil and grown under the same condition.

#### Vector construction and plant transformation

Gateway cloning technology (Invitrogen, USA) was used to generate *HsAlg10* and *HsAlg10B* expression vector for plants. *HsAlg10^WT^*, *HsAlg10^K^*^371f^*^s^*, *HsAlg10B^WT^*, and *HsALG10B^L^*^253^*^W^* were amplified from those in the pGW-HA-attB plasmids. The amplicons were inserted into pENTR™/D-TOPO™ entry vector (Invitrogen, USA). AtALG10 in pENTR™/SD/D-TOPO™ entry vector was acquired from ABRC. Gateway LR Clonase II Enzyme mix (Invitrogen, USA) was used to transfer the insert in the entry vector to the pK7WG2 destination vector ^83^. The expression of the insert was controlled by the CaMV35s constitutive promoter in the pK7WG2 vector. The recombinant plasmids were transformed to *alg10-1* mutant by a floral dip approach using *Agrobacterium tumefaciens* strain GV3101 ^110^. Transformants were grown on ½ MS agar plates supplemented with 50 µg/mL kanamycin (Fisher Scientific, USA) for three generations to select homozygous transformants. PCR was used to confirm the transformants. Seeds from the third-generation plants were used in the germination assay.

#### Arabidopsis Germination assay

Seeds of Col-0, *alg10-1*, and *alg10-1* containing *35s::AtALG10*, *35s::HsAlg10^WT^*, *35s::HsAlg10^K^*^371f^*^s^*, *35s::HsAlg10B^WT^*, and *35s::HsALG10B^L^*^253^*^W^*were sterilized as previously described. Seeds were plates on ½ MS agar plates supplemented with 100 and 150 mM NaCl and were stratified at 4°C for three days. Plates were placed vertically in the growth chamber with the same conditions as previously described. The number of germinating seeds was counted for seven consecutive days.

The R package ‘germinationmetrics’ was applied to calculate germination percentage, mean germination time, and germination index ^115^ using the equations below.

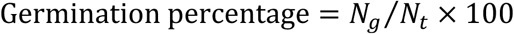

where

*N_g_* was the number of germinated seeds

*N_t_* was the total number of seeds ^116^

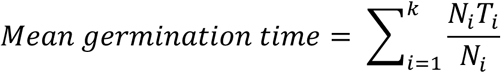

where

*T_i_* is the time from the beginning of the experiment to the ith interval,

*N_i_* is the number of seeds germinated on day

*k* is the total number of time intervals ^117^ ^118^

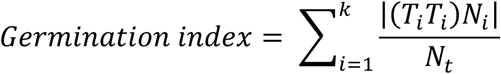

where

*T_i_* is the time from the beginning to the *i* th interval,

*N_i_* is the number of seeds germinated on day *i*,

*N_t_* is the total number of seeds, and

*k* is the total number of time intervals (Melville et al. 1980)

The germination assays were performed at least three times, and the results were an average from three independent experiments. Statistical analysis and plotting graphs were conducted in R version 4.0.3. One-way ANOVA with Šídák’s multiple comparisons test were performed using GraphPad Prism.

#### Arabidopsis Reverse Transcription Quantitative PCR (RT-qPCR)

Whole seedlings (14 days old) were flash-frozen in liquid nitrogen and stored at -80°C until use. Total RNA was extracted from plant tissues using RNeasy Plant Mini Kit (Qiagen, USA) with an addition of one nanogram of RNA spike (ArrayControl™ RNA Spikes, Invitrogen, USA) during mixing ground tissue in RTL buffer. The on-column DNase I treatment (Roche, USA) was performed to remove DNA. 2 µg of total RNA were used in cDNA synthesis (iScript™ Advance cDNA Synthesis Kit for RT-qPCR, Bio-Rad, USA). cDNAs were then 1:20 diluted in nuclease-free water for qPCR. iTaq™ Universal SYBR® Green Supermix (Bio-Rad, USA) was used for dye-based qPCR. One reaction (10 µl) consisted of 5 µL of SYBR® Green Supermix, 1 µL of 5 µM forward primer, 1 µL of 5 µM reverse primer, 2 µL of diluted cDNA, and 1 µL of nuclease-free water. The qPCR was performed on CFX384 Touch Real-Time PCR Detection System (Bio-Rad, USA) with the following conditions: initial denaturation at 95°C for 30 seconds and the 35 cycles of denaturation (95°C for 5 seconds) and annealing (60°C for 30 seconds). Melt curve analysis (65 to 95°C with a 0.5°C increment) was applied at the end of qPCR cycles. Four technical replicates were performed for each cDNA sample in each target gene. Only samples with a single discrete peak in the melt curve analysis were retained for gene expression analysis. Relative normalized expression (ΔΔCq) was determined by CFX Maestro 1.1 software (Bio-rad, USA) using *ACTIN 2* (*ACT2*), *ISOPENTENYL DIPHOSPHATE ISOMERASE 2* (*IPP2*), and ArrayControl RNA spikes as reference genes.

#### Arabidopsis protein extraction and immunoblot analysis

Total proteins were extracted from 14-day-old seedlings, and an extraction protocol was adapted from ^111^. After grinding tissues in liquid nitrogen, 500 µL of protein extraction buffer (1x PBS with protease inhibitor cocktail tablet (Roche, USA) was added to the ground tissues. Samples were incubated on ice for 10 minutes and centrifuged at 12,700 rpm for 10 minutes. The supernatant was collected and centrifuged again at the same speed for 5 minutes. The supernatant was collected as a protein crude extract. Samples were mixed with the SDS-PAGE loading buffer and heated at 95°C for 5 minutes. Samples were then subjected to the 7.5% SDS-PAGE under reducing conditions. Proteins on the gel were blotted onto the PVDF membrane using the Trans-Blot Turbo system (Bio-Rad, USA). The membrane was stained with Ponceau S solution for 10 minutes. For the immunoblot with anti-HRP, the membrane was incubated in the blocking buffer (1x TBST (20 mM Tris-HCl, 500 mM NaCl, 0.05% Tween-20; pH 7.5) with 5% nonfat dry milk) for 1 hour. After washing in 1x TBST, the membrane was incubated in the 1:2,000 mouse polyclonal anti-HRP antibody (Abcam, USA) overnight at 4°C. The 1:10,000 goat anti-mouse IgG antibody conjugated with IRDye® 800CW (Abcam, USA) was used to detect the binding of anti-HRP on the membrane. The membrane was incubated in the secondary antibody for 1 hour and then washed in 1x TBST. The fluorescence signal on the membrane was captured by Odyssey CLx Imager (Li-Cor, USA). For the lectin blot, 1x TBST with 3% bovine serum albumin was used as a blocking buffer. The membrane was incubated in the blocking buffer for 1 hour and then washed with 1x TBST. The membrane was incubated in the 1:5,000 biotinylated concanavalin A (Con A) (Vector laboratories, USA) overnight at room temperature and then washed with 1x TBST. The 1:20,000 IRDye® 800CW Streptavidin (Li-Cor, USA) was used to determine the biotinylated Con A by incubating the membrane in the solution for 1 hour. The membrane was washed with 1x TBST and captured by Odyssey CLx Imager (Li-Cor, USA).

### Proteomics and glycoproteomics

#### Global proteomic and N-glycoproteomic analysis

Flies of the following genotypes were cultured at 25°C in bottles: elav-Gal4>GFP^RNAi^, elav>Alg10^RNAi(S)^ and elav>Alg10^RNAi(VS)^. Adult flies 3 days post-eclosion from triplicate were decapitated were each done in triplicate and compared to a control (GFP) RNAi in quadruplicate. Proteins were extracted according to the method described by Hurkman and Tanaka with some modifications ^112^. Briefly, proteins were isolated by phenol and precipitated overnight by five volumes of pre-chilled 0.1 M ammonium acetate in 100% methanol. Brain samples were lysed in 1 mL of lysis buffer (20 mM HEPES pH 7.9, 1% SDS, and protease inhibitors) with sonication. Protein concentrations were determined by BCA assay (Thermo Fisher Scientific, Cat# 23225). Reduction and alkylation were performed as previously described ^113^. S-trap digestion was performed according to the manufacturer’s instructions (ProtiFi, Cat# C02-mini), resulting in ∼600 µg tryptic peptides per sample (300 μL).

To half of the tryptic peptides, 15 mg of HILIC beads (PolyLC, Cat# BMHY0502) pre-activated with 0.1% trifluoroacetic acid (TFA) were added to yield a 1:50 peptide-to-beads mass ratio. The samples were vortexed in binding buffer (0.1% TFA, 19.9% H_2_O, 80% ACN) for 1 h at 24 °C to allow *N*-glycopeptides to bind to the beads. The unbound peptides were removed by washing the beads with 6 x 150 μL binding buffer. *N*-glycopeptides were eluted from the beads with 5 x 150 μL of 0.1% TFA/99.9% H2O. Finally, 2 μL PNGase F (500 U/μL) (NEB # P0704) was added to the elution buffer and the samples were incubated for 3 h at 37 °C. The eluted *N*-glycopeptides were concentrated to dryness.

To the other half of the tryptic peptides, multi-lectin enrichment was performed based on the previously reported protocol by Mann and co-workers with some modifications ^114^. Briefly, tryptic peptides were treated with a mixture of lectins 90 μg ConA, 90 μg WGA, and 36 μg RCA120 (Sigma-Aldrich, Cat# L7886, Cat# L9640, and Cat# L7647) in 2 × binding buffer and transferred to a 30 kDa filter (Pall Corporation, Cat# OD030C34). The mixture was incubated for 1 h at 24 °C and unbound peptides were removed by centrifugation at 14 000 × g for 10 min. The remaining *N*-glycopeptides bound to lectins were washed with 200 μL binding solution for four times and 50 μL digest buffer of 50 mM citrate-phosphate buffer (Sigma-Aldrich, Cat# P4809) twice. Finally, 2 μL PNGase F (5U/μL) (NEB # P0704S) was added to the filter and incubated for 3 h at 37 °C. The deglycosylated *N*-glycopeptides were eluted with 2 × 50 μL digest buffer.

N-Glycopeptides from both enrichment methods were combined and desalted by C18 Tips (Thermo Fisher Scientific, Cat# 87784) following the manufacturer’s instructions. The desalted peptides were resuspended in 30 µL 100 mM triethylammonium bicarbonate (TEAB) buffer. For each sample, 5 µL of the corresponding amine-based tandem mass tag (TMT) 10-plex reagent (10 µg/µL) was added and reacted for 1 h at 24 °C. The reactions were quenched with 2 µL 5% hydroxylamine solution (Oakwood Chemical, Cat# 069272) and the combined mixture was concentrated to dryness. High-pH fractionation was performed according to the manufacturer’s instructions (Thermo Fisher Scientific, Cat# 84868), which resulted in eight fractions. For the global proteomic analysis, ∼0.3 mg tryptic peptides were reacted with 20 µL of TMT 10-plex reagents and fractionated into 20 samples. All samples were stored at –80 °C until further analysis.

#### Mass spectrometry acquisition procedures

A Thermo Scientific EASY-nLC 1000 system was coupled to a Thermo Scientific Orbitrap Fusion Tribrid with a nano-electrospray ion source. Mobile phases A and B were water with 0.1% formic acid (v/v) and acetonitrile with 0.1% formic acid (v/v), respectively. For each fraction, peptides were separated with a linear gradient from 4% to 32% B within 50 min, followed by an increase to 50% B within 10 min and further to 98% B within 10 min, and re-equilibration. The instrument parameters were set as follows: survey scans of peptide precursors were performed at 120K FWHM resolution over a m/z range of 410– 1800. HCD fragmentation was performed on the top 10 most abundant precursors exhibiting a charge state from 2 to 5 at a resolving power setting of 50K and fragmentation energy of 37% in the Orbitrap. CID fragmentation was applied with 35% collision energy and resulting fragments detected using the normal scan rate in the ion trap.

#### Mass spectrometry data analysis

The raw data was processed using Proteome Discoverer 2.4 (Thermo Fisher Scientific). Data was searched against the UniProtKB Fruit fly (Drosophila melanogaster) protein database (Nov. 2019, 21,041 total entries) and common contaminant proteins using Sequest HT and Byonic algorithms. Searches were performed with the following guidelines: spectra with a signal-to-noise ratio greater than 1.5; trypsin as enzyme, 2 missed cleavages; variable oxidation on methionine residues (15.995 Da) and deamidation on asparagine (0.984 Da); static carboxyamidomethylation of cysteine residues (57.021 Da), static TMT labeling (229.163 Da) at lysine residues and peptide N-termini; 10 ppm mass error tolerance on precursor ions, and 0.02 Da mass error on fragment ions. Data were filtered with a peptide-to-spectrum match (PSM) of 1% FDR using Percolator. The TMT reporter ions were quantified using the Reporter Ions Quantifier without normalization. For the global proteomic analysis, the obtained PSMs were filtered with the following guidelines: confidence is high, PSM ambiguity is unambiguous, unique peptides are greater than two and exclude all contaminant proteins. Data analysis was performed as previously described ^119^. For the *N*-glycoproteomic analysis, the obtained PSMs were filtered with the following guidelines: confidence is high, PSM ambiguity is unambiguous, modifications contain deamidated and exclude all contaminant proteins. Data analysis was performed by an in-house script. Briefly, consensus sequence filtering (N-X-S/T) was applied using a string match module. The generation of missing values and VSN normalization were performed as previously described ^120^. Fold Change (FC) was presented after generalized log transformation and normalization of the raw ratio between genotypes to fit a linear model, with log_2_FC values equaling +/– 0.73, corresponding to a raw fold change > 2.

All analyses were performed in R. UniProt IDs were mapped to fly genes. Comparisons were made using T-test and correction for multiple testing was based on FDR thresholds. Analyses were performed both at the gene (protein) level, i.e., combining multiple glycosylation sites in a single protein-coding gene, as well as the level of individual glycosites. Effect size was calculated using fold changes in the median levels across groups.

We retained only those peptides that could be unambiguously matched to a unique protein sequence and analyzed the glycoproteome at the glycosite level and at the glycoprotein level (where multiple glycosites in a protein are collapsed into a single number).

## ACKNOWLEDGEMENTS

We would like to thank members of our labs for critical comments on the manuscript. We acknowledge funding from the following sources:

T.M.: NIH T32HL007901.

S.T.: Banting and NSERC postdoctoral fellowships.

K.L.: Development and Promotion of Science and Technology Talents Project (DPST), Thailand.

J.M.L.: NIH R01 HG012810 and R35 GM146839.

K.G.O.: NIH R15GM155985.

B.M.: NIH T32MH020068 and T32NS062443.

C.E.: HHMI Hanna Gray Fellowship.

B.dB.: NIH-NINDS 1R01NS121874-01.

J.T.L.: NIH MH104536 and NS117588.

C.J.D.: USDA National Institute of Food and Agriculture Project 1002035 and National Science Foundation (NSF) 2210293.

G.C.M.: NIH-NIA 1R01AG065992.

D.A.P.: NIH R35 NS122172.

C.M.W. and J.A.W.: NIH 1RF1AG081475-01.

R.S., J.A.W., G.C.M.: NIH-NHLBI 1-R01 HL146751-01A1.

*Drosophila* stocks obtained from the Bloomington Drosophila Stock Center (NIH P40OD018537) were used in this study. Some *C. elegans* strains were provided by the *Caenorhabditis* Genetics Center (CGC), which is funded by NIH Office of Research Infrastructure Programs (P40 OD010440). We thank both Flybase and WormBase.

## AUTHOR CONTRIBUTIONS

S.G., T.R.M, S.L.S., R.S., J.A.W. conceived the idea and designed the experiments.

J.A.W., R.S., S.L.S. provided overall project leadership and funding support.

J.M.L., M.M., S.G. and R.S. performed the human genetic analyses.

S.G., T.R.M, B.L., S.R.M., N.L., S.S. and J.A.W. performed the *Drosophila* genetics and behavioral experiments.

O.K. and H.J.B. designed and generated the *Drosophila* transgenic constructs.

K.G.O. and J.T.L. performed the *Drosophila* electrophysiology experiments.

C.E, J.A-Z. and B.dB. designed and constructed the MARGO apparatus.

S.T and D.A.P. carried out zebrafish experiments.

K.L. and C.J.D. designed and performed the *Arabidopsis* experiments.

A.C.H. designed *C. elegans* experiments carried out by B.M. and K.N.S. B.Y., C.A. and C.M.W. carried out proteomics experiments.

D.B. and G.C.M. performed the *Drosophila* cardiac assays.

A-E.L. and C.C. performed genotyping of human subjects.

J.W.W. assisted with patient diagnosis.

A.D.E., B.B., N.B., D.K. and G.B.S. have been providing standard of care for the PME proband, provided patient history and assessed sleep (PSG and actigraphy) and brain structure (MRIs) for this study.

S.G., T.R.M., R.S. and J.A.W. wrote the paper.

## DECLARATION OF INTERESTS

S.G. is a stockholder and employee of Magnet Biomedicine. S.L.S. and R.S. are founders and stockholders of Magnet Biomedicine. B.Y. is a stockholder and employee of Sanofi. The other authors declare no competing interests.

## Notes

### Funding Statement

R.S., J.A.W., G.C.M.: NIH-NHLBI 1R01 HL146751-01A1.
C.M.W. and J.A.W.: NIH 1RF1AG081475-01
G.C.M.: NIH-NIA 1R01AG065992.
B.dB.: NIH-NINDS 1R01NS121874-01.
J.T.L.: NIH MH104536 and NS117588.
D.A.P.: NIH R35 NS122172.
C.J.D.: USDA National Institute of Food and Agriculture Project 1002035 and National Science Foundation (NSF) 2210293
J.M.L.: NIH R01 HG012810 and R35 GM146839.
K.G.O.: NIH R15GM155985
B.M.: NIH T32MH020068 and T32NS062443.
No authors or their institutions received payment or services from a third party for any aspect of the submitted work at any time.

### Author Declarations

Ethics committee of İSTANBUL UNIVERSITY, Deanery of İstanbul Medical Faculty gave ethical approval for this work Date: 11/06/2021 (Number: E-29624016-050.99-267817, Topic: Regarding Prof. Dr. Betul Baykan BAYKAL, Evaluation of Epilepsy and Sleep Disorders in Patients with ALG10 mutations filed under 2021/1045)

