## Supplemental Figures for "A conserved role for ALG10/ALG10B and the *N*-glycosylation pathway in the sleep-epilepsy axis"

Fig. S1

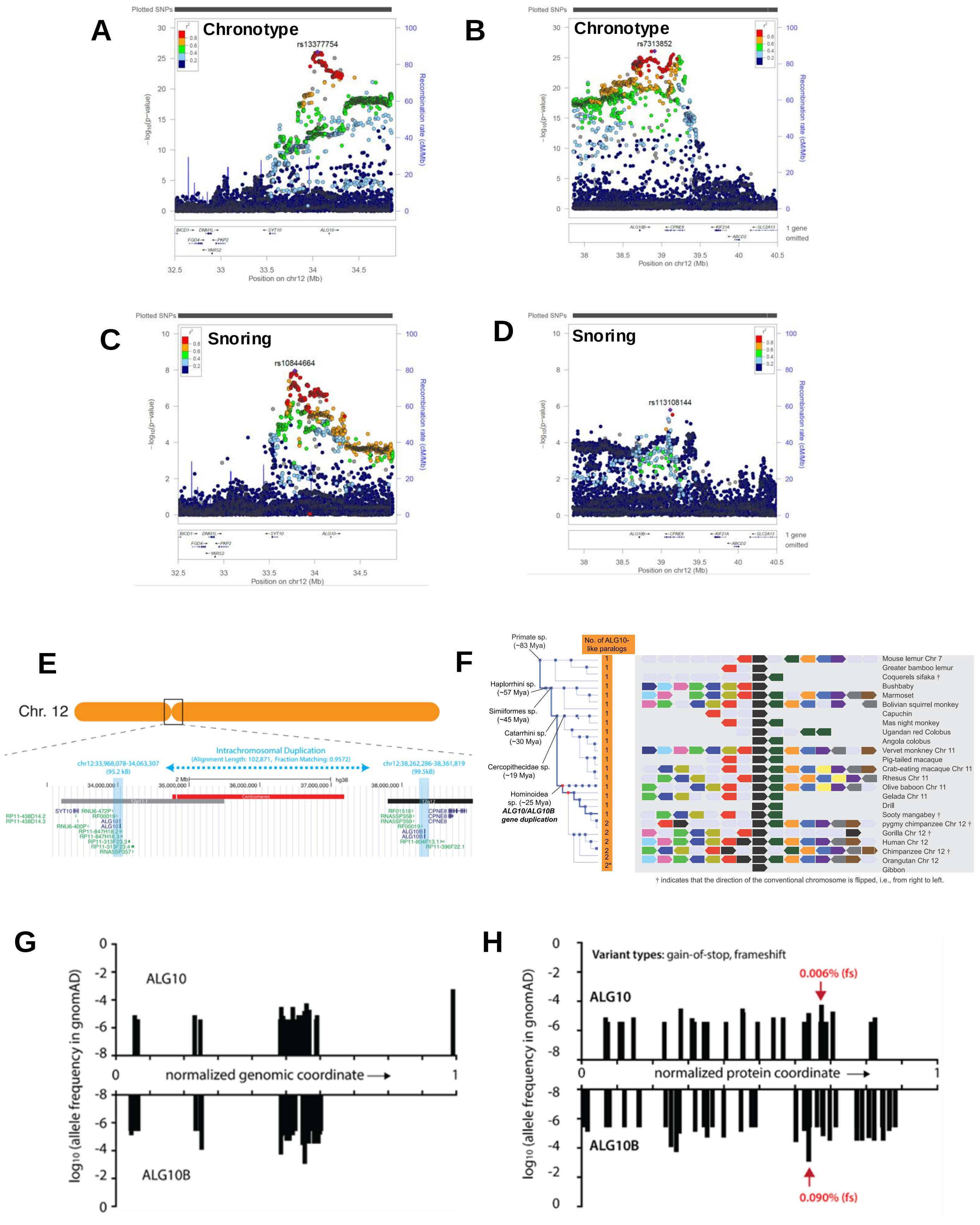

A
Chronotype
rs13377754
Plotted SNPs
-log10(p-value)
Recombination rate (cM/Mb)
Position on chr12 (Mb)
B
Chronotype
rs7313852
1 gene omitted
C
Snoring
rs10844664
D
Snoring
rs113108144
E
Chr. 12
Intrachromosomal Duplication
(Alignment Length: 102,871, Fraction Matching: 0.9572)
chr12:33,968,078-34,063,307
(95.2 kB)
chr12:38,262,286-38,361,819
(99.5kB)
Centromeres
F
No. of ALG10-like paralogs
Primate sp. (~83 Mya)
Haplorrhini sp. (~57 Mya)
Simiiformes sp. (~45 Mya)
Catarrhini sp. (~30 Mya)
Cercopithecidae sp. (~19 Mya)
Hominoidea sp. (~25 Mya)
ALG10/ALG10B gene duplication
Mouse lemur Chr 7
Greater bamboo lemur
Coquerels sifaka †
Bushbaby
Marmoset
Bolivian squirrel monkey
Capuchin
Mas night monkey
Ugandan red Colobus
Angola colobus
Vervet monkney Chr 11
Pig-tailed macaque
Crab-eating macaque Chr 11
Rhesus Chr 11
Olive baboon Chr 11
Gelada Chr 11
Drill
Sooty mangabey †
pygmy chimpanzee Chr 12 †
Gorilla Chr 12 †
Human Chr 12
Chimpanzee Chr 12 †
Orangutan Chr 12
Gibbon
† indicates that the direction of the conventional chromosome is flipped, i.e., from right to left.
G
ALG10
ALG10B
normalized genomic coordinate
log10 (allele frequency in gnomAD)
H
Variant types: gain-of-stop, frameshift
ALG10
ALG10B
0.006% (fs)
0.090% (fs)
normalized protein coordinate
log10 (allele frequency in gnomAD)

### Fig. S2

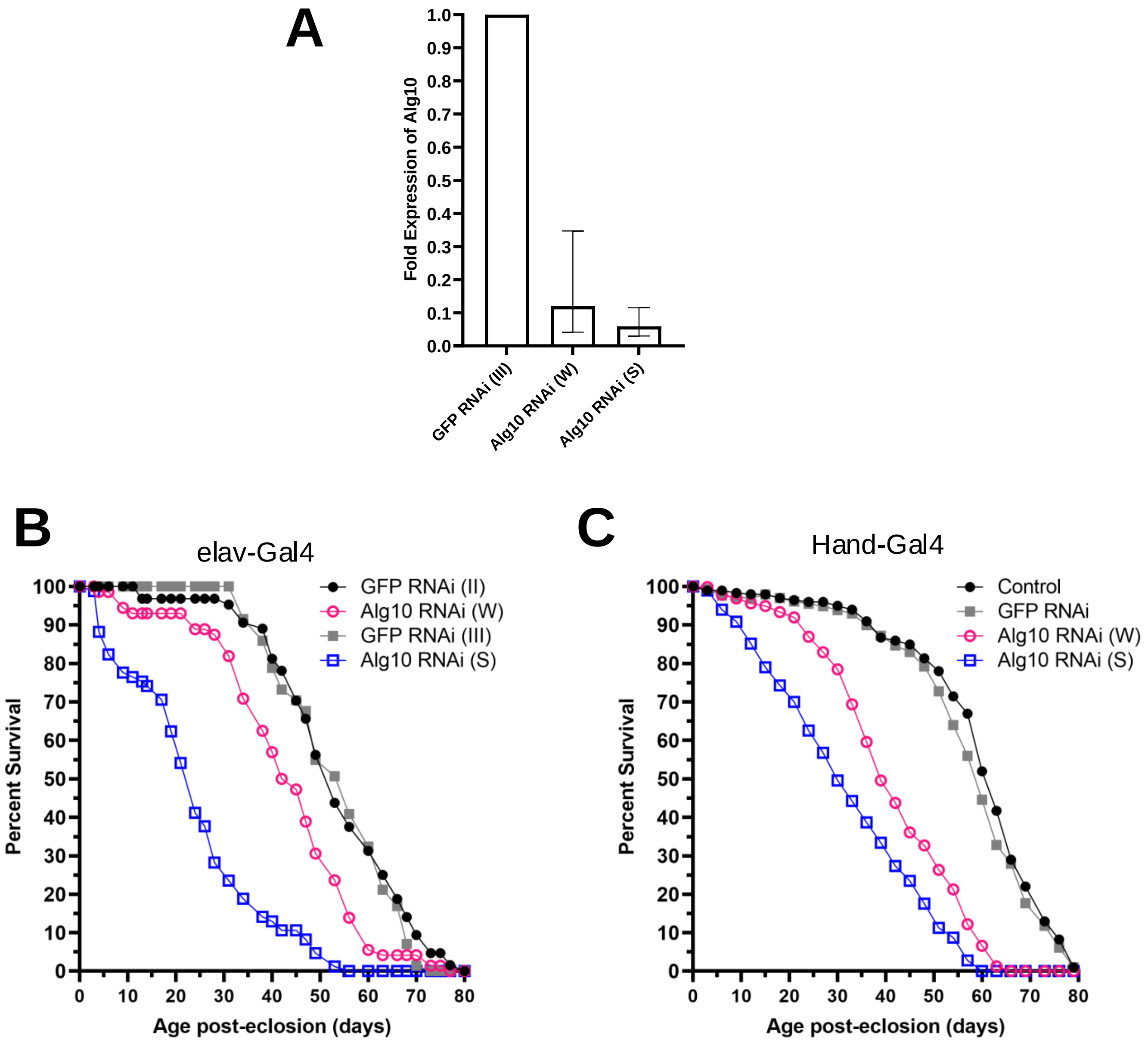
A
Fold Expression of Alg10
1.0
0.9
0.8
0.7
0.6
0.5
0.4
0.3
0.2
0.1
0.0
GFP RNAi (III)
Alg10 RNAi (W)
Alg10 RNAi (S)
B
elav-Gal4
GFP RNAi (II)
Alg10 RNAi (W)
GFP RNAi (III)
Alg10 RNAi (S)
Percent Survival
Age post-eclosion (days)
C
Hand-Gal4
Control
GFP RNAi
Alg10 RNAi (W)
Alg10 RNAi (S)
Percent Survival
Age post-eclosion (days)

### Fig. S3

## A

| Driver | RNAi | N Flies | Percent Rhythmic | Rhythmic | Arrhythmic | Rhythmic Only | | | | | |
|---|---|---|---|---|---|---|---|---|---|---|---|
| | | | | | | Avg FFT | St. Dev. | Adj. P Value | Avg Period | St. Dev. | Adj. P Value |
| ElaV | GFP RNAi III | 56 | 83.9% | 47 | 9 | 0.0111 | 0.0041 | 0.0211 | 23.64 | 1.06 | 0.9999 |
| | Alg10 RNAi (S) | 53 | 77.4% | 41 | 12 | 0.0091 | 0.0035 | | 23.65 | 0.42 | |
| Cry | GFP RNAi III | 20 | 90.0% | 18 | 2 | 0.0128 | 0.0037 | 0.1681 | 25.11 | 1.25 | 0.8704 |
| | Alg10 RNAi (S) | 38 | 81.6% | 31 | 7 | 0.0110 | 0.0025 | | 24.96 | 1.53 | |

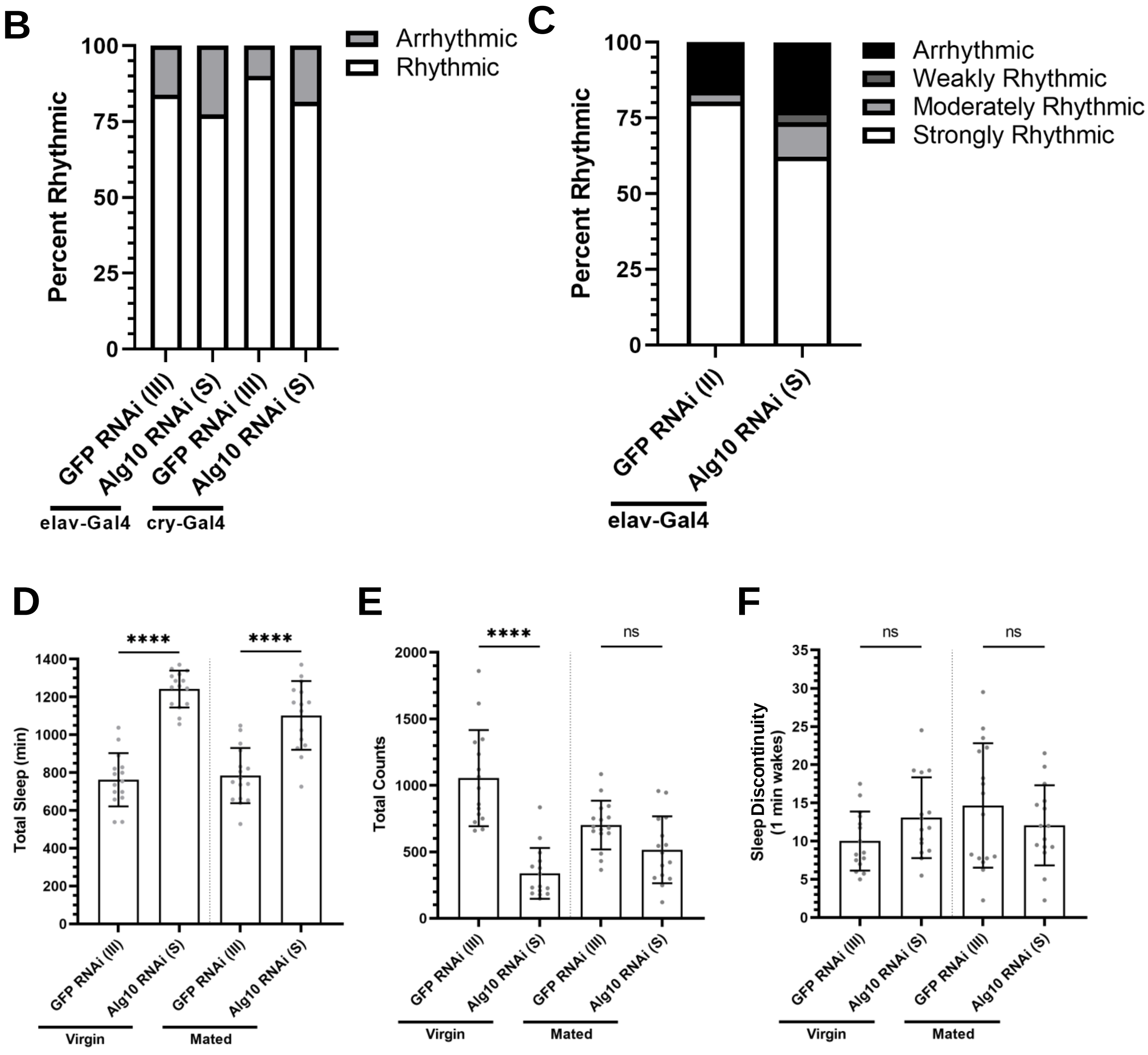

Fig. S4

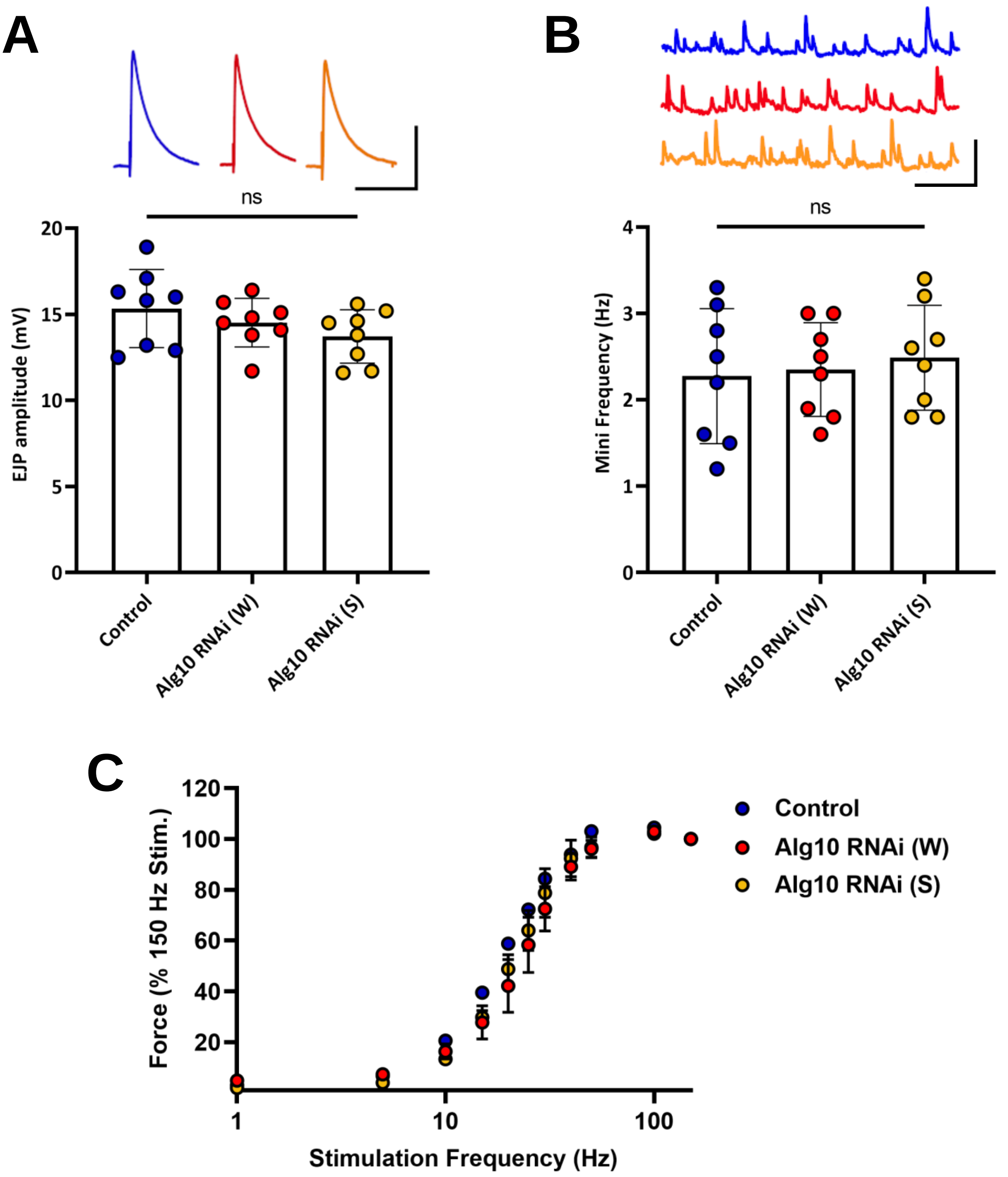

A
ns
EJP amplitude (mV)
20
15
10
5
0
Control
Alg10 RNAi (W)
Alg10 RNAi (S)
B
ns
Mini Frequency (Hz)
4
3
2
1
0
Control
Alg10 RNAi (W)
Alg10 RNAi (S)
C
Force (% 150 Hz Stim.)
120
100
80
60
40
20
1
10
100
Stimulation Frequency (Hz)
Control
Alg10 RNAi (W)
Alg10 RNAi (S)

### Fig. S5

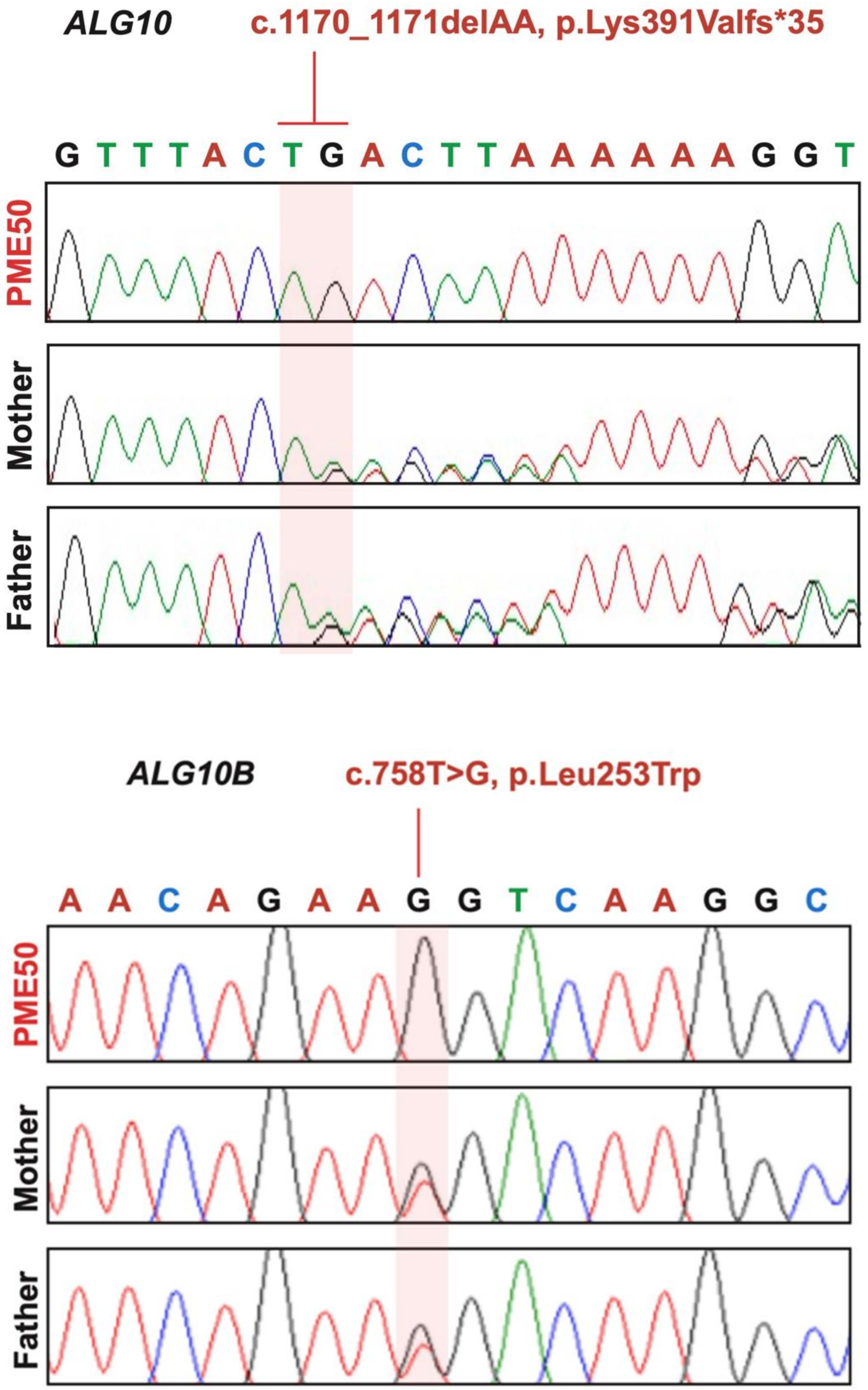

ALG10
c.1170_1171delAA, p.Lys391Valfs*35
G T T T A C T G A C T T A A A A A A G G T
PME50
Mother
Father
ALG10B
c.758T>G, p.Leu253Trp
A A C A G A A G G T C A A G G C
PME50
Mother
Father

### Fig. S6

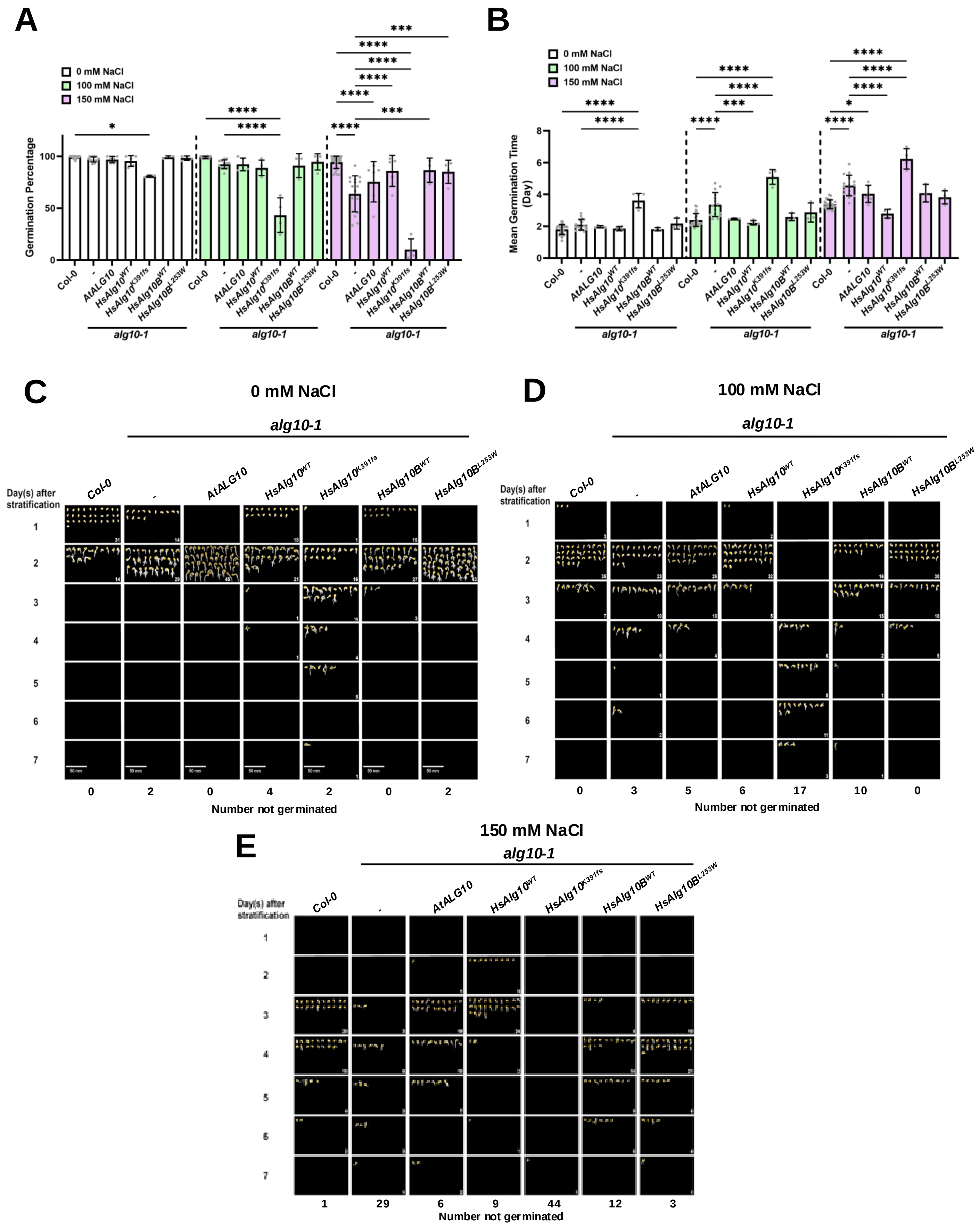
A
0 mM NaCl
100 mM NaCl
150 mM NaCl
Germination Percentage
0
50
100
Col-0
-
AtALG10
HsAlg10WT
HsAlg10K391fs
HsAlg10BWT
HsAlg10BL253W
alg10-1
B
Mean Germination Time (Day)
0
2
4
6
8
C
0 mM NaCl
alg10-1
Day(s) after stratification
1
2
3
4
5
6
7
50 mm
0
2
0
4
2
0
2
Number not germinated
D
100 mM NaCl
alg10-1
0
3
5
6
17
10
0
Number not germinated
E
150 mM NaCl
alg10-1
1
29
6
9
44
12
3
Number not germinated

### Fig. S7

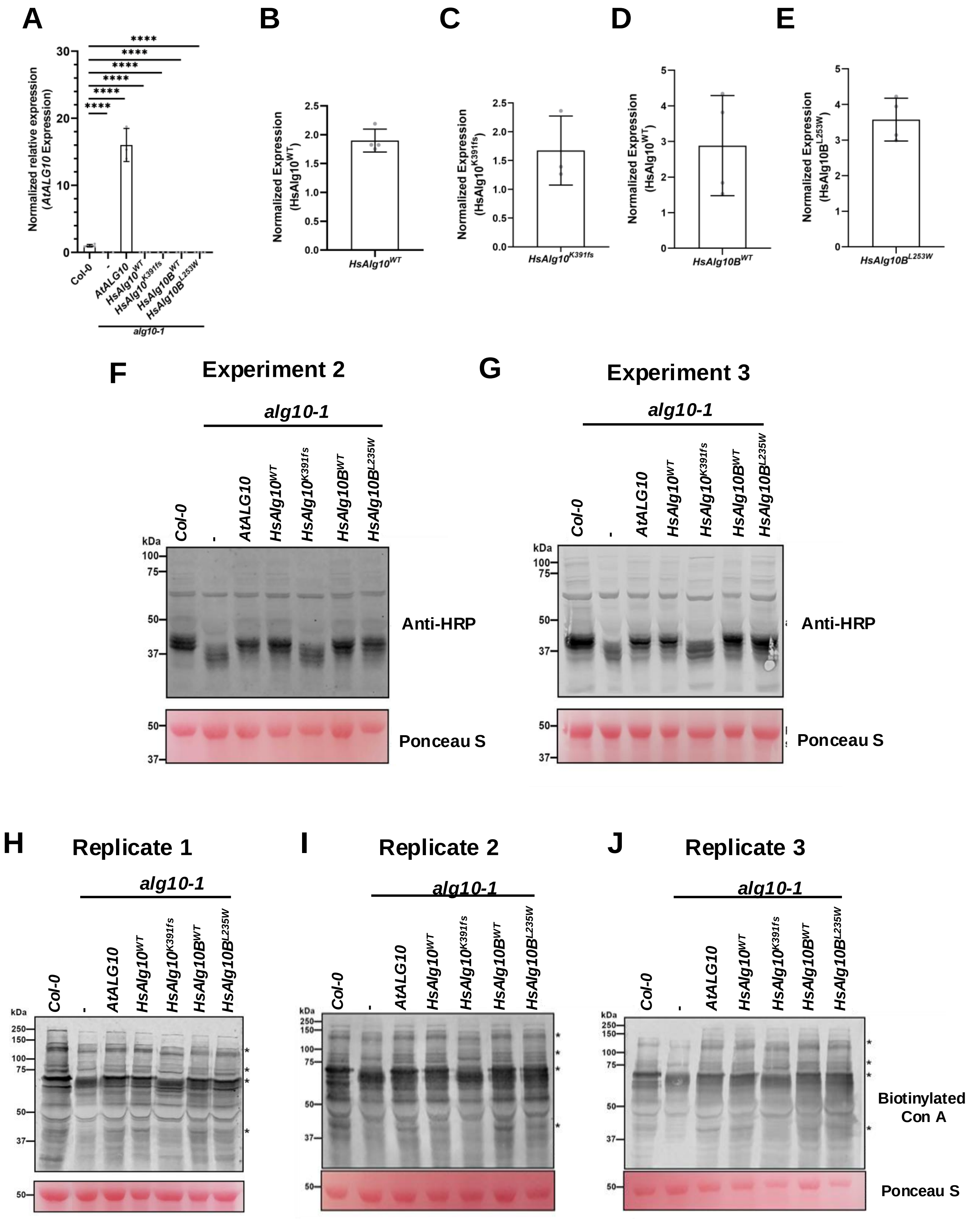

A
B
C
D
E
Normalized relative expression (AtALG10 Expression)
Col-0
-
AtALG10
HsAlg10WT
HsAlg10K391fs
HsAlg10BWT
HsAlg10BL253W
alg10-1
Normalized Expression (HsAlg10WT)
Normalized Expression (HsAlg10K391fs)
Normalized Expression (HsAlg10BWT)
Normalized Expression (HsAlg10BL253W)
F
Experiment 2
G
Experiment 3
HsAlg10K391fs
HsAlg10BL235W
kDa
Anti-HRP
Ponceau S
H
Replicate 1
I
Replicate 2
J
Replicate 3
Biotinylated Con A

### Fig. S8

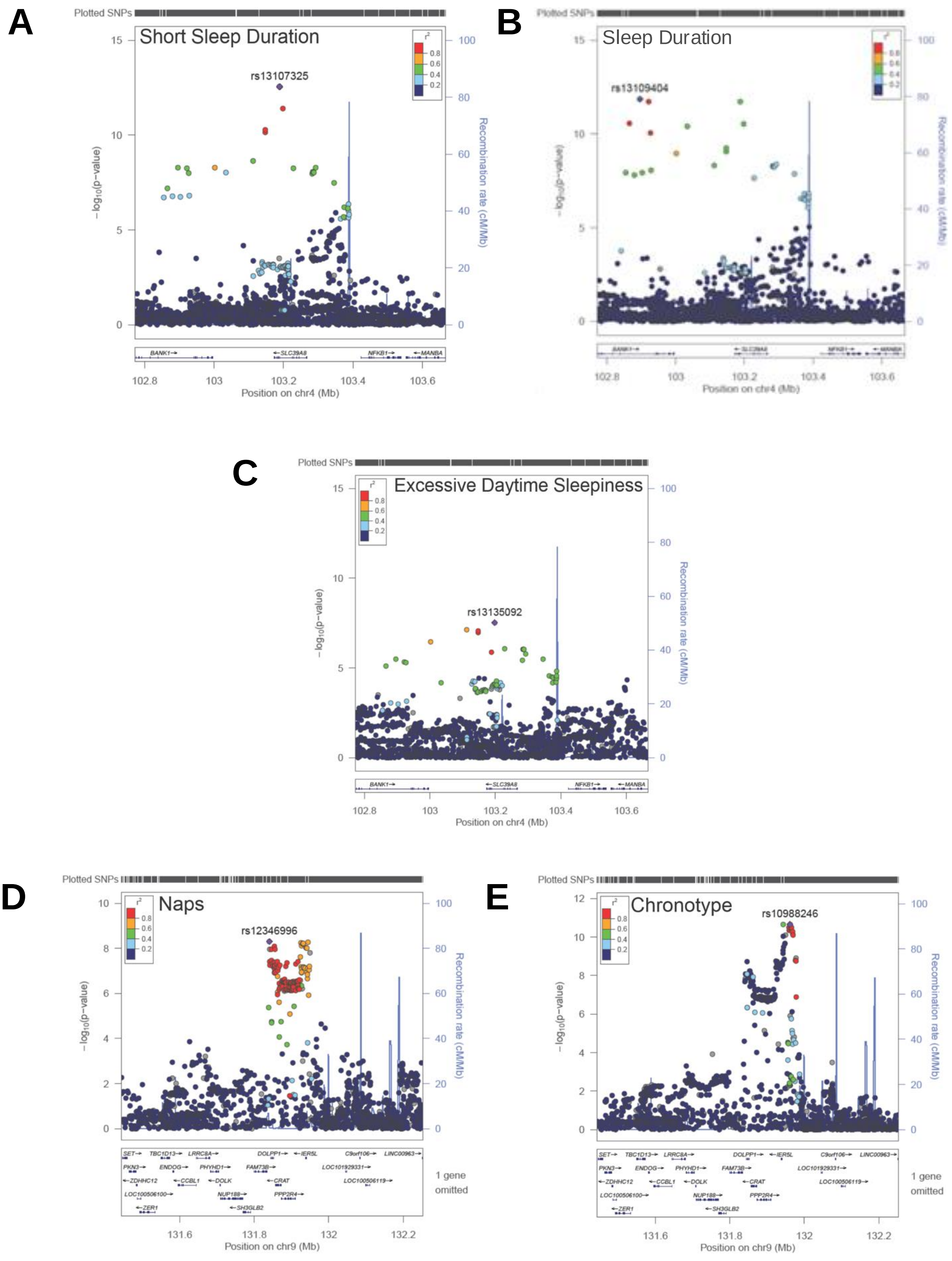
A
Short Sleep Duration
rs13107325
Position on chr4 (Mb)
B
Sleep Duration
rs13109404
Position on chr4 (Mb)
C
Excessive Daytime Sleepiness
rs13135092
Position on chr4 (Mb)
D
Naps
rs12346996
Position on chr9 (Mb)
E
Chronotype
rs10988246
Position on chr9 (Mb)
